# ^1^H-NMR metabolomics-guided DNA methylation mortality predictors

**DOI:** 10.1101/2023.11.02.23297956

**Authors:** D. Bizzarri, M.J.T. Reinders, L.M. Kuiper, M. Beekman, J. Deelen, J.B.J. van Meurs, J. van Dongen, R. Pool, D.I. Boomsma, M. Ghanbari, L. Franke, BIOS Consortium, BBMRI-NL Consortium, P.E. Slagboom, E.B. van den Akker

## Abstract

^1^H-NMR metabolomics and DNA methylation in blood are widely known biomarkers predicting age-related physiological decline and mortality yet exert mutually independent mortality and frailty signals. Leveraging multi-omics data in four Dutch population studies (N=5238) we investigated whether the mortality signal captured by ^1^H-NMR metabolomics could guide the construction of novel DNA methylation-based mortality predictors. Hence, we trained DNA methylation-based surrogates for 64 metabolomic analytes and found that analytes marking inflammation, fluid balance, or HDL/VLDL metabolism could be accurately reconstructed using DNA-methylation assays. Interestingly, a previously reported multi-analyte score indicating mortality risk (MetaboHealth) could also be accurately reconstructed. Sixteen of our derived surrogates, including the MetaboHealth surrogate, showed significant associations with mortality, independent of other relevant covariates. Finally, adding our novel surrogates to previously established DNA-methylation markers, such as GrimAge, showed significant improvement for predicting all-cause mortality, indicating that our metabolic analyte-derived surrogates potentially represent novel mortality signal.

## Introduction

A common goal in geroscience is to identify mechanisms that drive ageing and design interventions that might slow down or even reverse the rate of ageing [1]. For this purpose, it is essential to have indicators not only quantifying ageing, but simultaneously marking the trajectory of overall health decline [2]. While calendar age is a core risk factor for almost any common disease, it has many limitations for capturing the variability in health-span. Crucially, calendar age does not capture the effects of an individual’s lifestyle, nor incorporates readouts of functional decline. Instead, faithful markers of biological age would allow to quantify the vulnerability to disease irrespective of an individual’s calendar age, and to develop and monitore effective healthy lifestyle advices and anti-aging interventions. The earliest approaches to construct such markers of biological age relied on clinical measures of physiological capacity [3]. Later molecular and -omics approaches gained popularity, initially including markers such as leukocyte telomere length [4], followed by multi-marker algorithms based on high-throughput platforms, such as DNA methylation [5], transcriptomics [6], metabolomics [7], and proteomics [8]. Importantly, these algorithms were trained to estimate cross-sectional chronological age. Of these omics approaches, particularly DNA methylation-based algorithms exhibited remarkably high accuracies in predicting calendar age [5], and were named ‘DNA methylation clocks’. Nonetheless, while interesting by itself, this observation highlighted a fundamental limitation in this first design of markers of biological age. Since nearly-perfect age predictors would arrive to similar observations as chronological age, they would lose their characteristics as age-independent health status indicators [9].

Concomitantly, a second generation of -omics markers was introduced, which instead were trained to predict the mortality risk. Prominent examples of these mortality-trained multivariate markers include the DNA methylation-based PhenoAge [10] and GrimAge [11] and the ^1^H-NMR metabolomics-based MetaboHealth [12]. These predictors were trained quite differently. The wide availability of the Nightingale Health ^1^H-NMR metabolomics in large prospective population studies, in combination with its relatively narrow though informative content (∼250 analytes), allowed for a more classic and direct approach. Deelen *et al.* trained MetaboHealth as a linear combination of 14 metabolic features, showing a strong predictive value, not only for mortality risk, but also for other outcomes, including pneumonia [13], and frailty [14]. Conversely, the DNA methylation platform by Illumina contains hundreds of thousands of features, and thus requires additional guidance to robustly capture the mortality signal. Hence, the PhenoAge and GrimAge were trained using the so-called two-stage approaches, in which more widely-available markers associated with mortality were leveraged to help extract the mortality signal [10,11]. PhenoAge achieved this by first training an all-cause mortality predictor based on clinical measures (e.g., glucose, C-reactive-protein), which was then re-estimated using DNA methylation. Similarly, DNAm-GrimAge is composed by a combination of DNA methylation-based surrogates for molecular or phenotypic markers known to associate with mortality. Interestingly, both two-step training strategies yielded DNA methylation-based scores that can associate not only with mortality, but also with a wide diversity of disease outcomes. These developments indicate that mortality-trained predictors for biological age can be trained using different omics platforms, and moreover, that DNA-methylation might serve as a platform to integrate these signals captured by different data sources. This latter concept was recently further substantiated by the work of Gadd *et al*., who systematically trained DNA-methylation-based predictors for 109 plasma proteins showing significant associations with incident morbidities over 14-years [15].

In a recent study we demonstrated that mortality-based predictors such as MetaboHealth and GrimAge are instrumental in predicting frailty in studies of middle-aged and elderly individuals [14]. Importantly, we also showed that these scores confer mutually independent information for predicting both frailty and mortality. Viewing these developments in the field, we thus pose the question to what extent the mortality signal captured by ^1^H-NMR metabolomics could be transferred and integrated with the mortality signals captured by the DNA-methylation platform. For this purpose, we will evaluate both strategies for training two-stage DNA-methylation based mortality predictors. On one hand, we will train a DNA methylation-based predictor re-estimating directly MetaboHealth, akin the strategy of PhenoAge. On the other hand, we will train DNA-methylation surrogates for single metabolomics features, and combine these in an overall score, akin GrimAge. Moreover, we will evaluate to what extent DNA-methylation surrogates features from different origins capture mutually independent signals, also with respect to predicting mortality risk.

## Results

### Cross-cohort calibration of ^1^H-NMR metabolomics data

To derive DNA methylation-based models predicting metabolic features we analyzed data gathered by partners of the BIOS consortium [16,17], totalling 4,334 individuals for whom both DNA methylation (Illumina 450k) and ^1^H-NMR Metabolomics (Nightingale Health Plc) data have been assayed. The resulting dataset had contributions of four independently collected population studies: LIFELINES-DEEP (LIFELINES), Leiden-Longevity-Study Partners-Offspring (LLS-PAROFFS), Rotterdam-Study (RS), and Netherlands-Twin-Register (NTR), each with their own inclusion criteria, as reflected by differences in subject characteristics that range from the younger and leaner population of NTR (mean age=37.57 years and mean BMI=24.32 cm/kg^2^) to the older and heavier population of RS (mean age=67.15 years and mean BMI=27,71 cm/kg^2^) (Figure 1, Supplementary Table S1,). A reduced dimensionality projection using a t-distributed neighbour embedding (tSNE) suggested that the interindividual variance in metabolomics data could not only be attributed to interindividual phenotypic variability but was also capturing some systematic differences between studies (Figure S3A-C). Following Makinen *et al.,* we implemented a calibration technique suitable for cross-cohort harmonization, which starts with the assumption that individuals with similar phenotypic characteristics should on average exhibit similar metabolomics profiles [18]. For this purpose, we identified pairs of samples across cohorts with matching age, sex, and BMI, and used LIFELINES as a common reference to calibrate the other studies (Figure S2, more details in **methods**). A t-SNE projection of the calibrated data revealed a substantial reduction of the systematic differences between studies, as also quantified by k-BET (k-nearest neighbor Batch Effect Test) (Figure 2A-B, S3). Principal Variance Component Analyses (PVCA) further confirmed this observation indicating that the variation attributable to study differences was attenuated, while maintaining the variation attributed to relevant biologically variability (Figure 2C).

**Figure 1:**
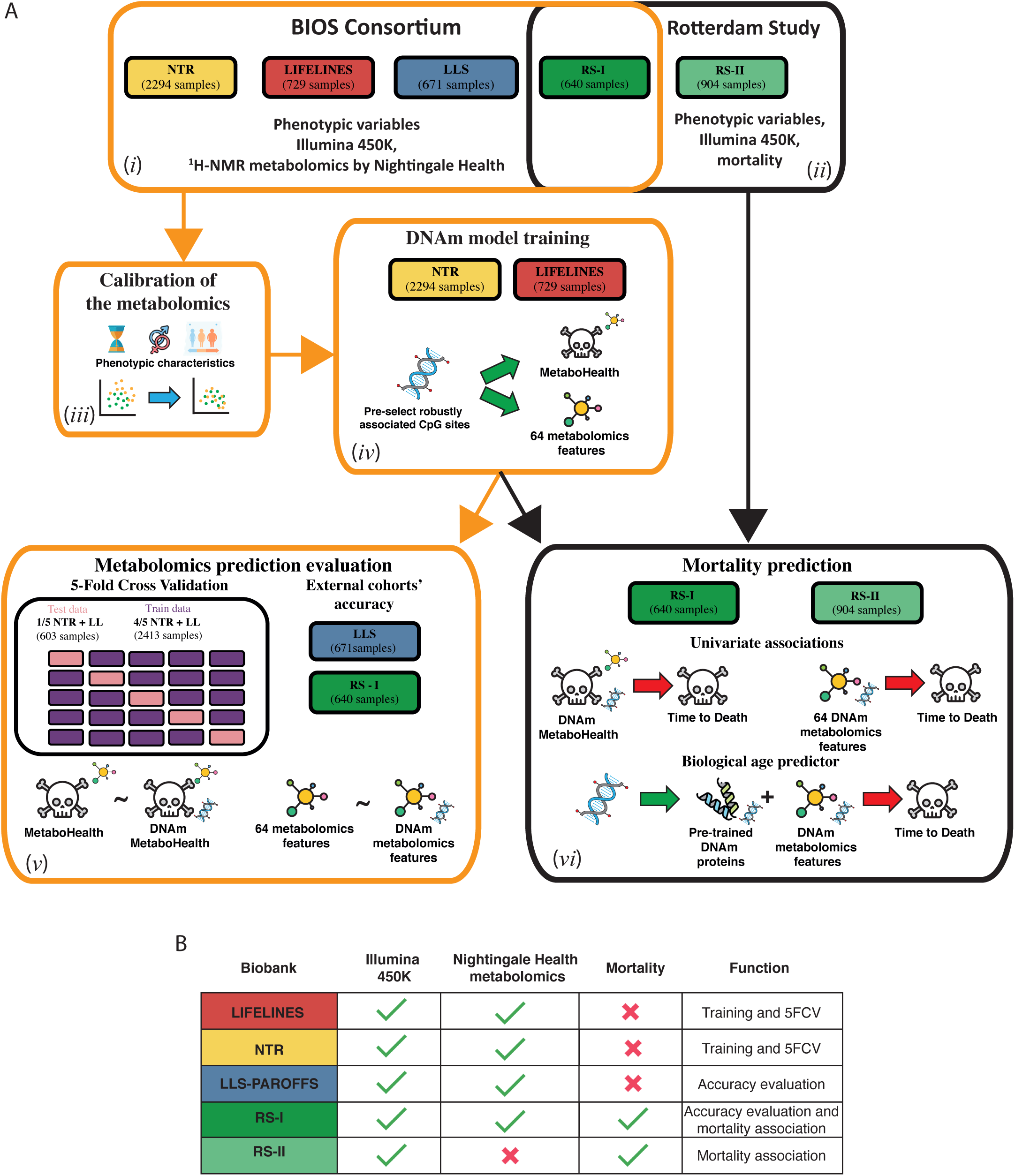
Study and methods overview. A) Study overview. (*i*) We employed 4334 samples, from 4 cohort of the BIOS Consortium, DNAm methylation and metabolomics to train and test our surrogates. (*ii*) Coupled with 1,544 samples from the Rotterdam Study to evaluate their associations with mortality. (*iii*) We applied a calibration to harmonize the metabolomics dataset. (*iv*) We then train ElasticNET models, on LIFELINES and NTR. Using the DNA methylation data we predict two types of outcomes: 1) the pre-trained metabolomics mortality predictor (MetaboHealth), and 2) the 64 metabolic features. (*v*) The DNAm models are evaluated using 1) the hold-out valuation sets (LLS and RS) and 2) a 5-Fold Cross Validation on the training sets (NTR and LIFELINES). (*vi*) Finally, we use the DNAm models to generate surrogate metabolomics features in the RS dataset (1544 samples) and 1) evaluate their univariate associations to mortality (while correcting for age, sex), and 2) trained a complete Cox regression combining our DNAm metabolomics features and the pre-trained DNAm surrogates. B) Availability of data in each cohort and when they are exploited in the study.

**Figure 2:**
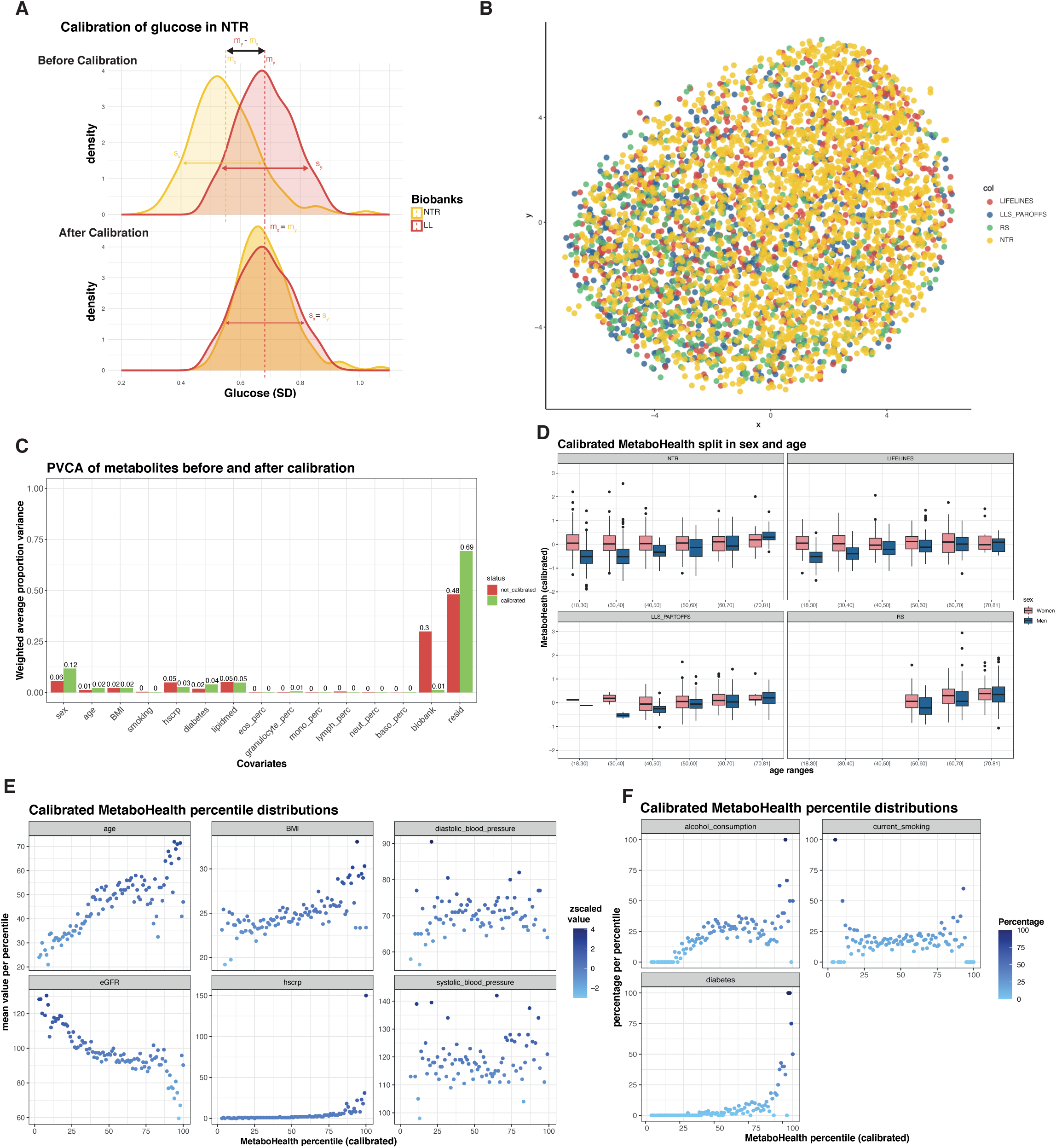
Harmonization of the metabolomics data. A) Distribution of glucose in NTR and LIFELINES before (upper figure) and after (lower figure) calibration. B) tSNE of the metabolomics dataset after calibration and colored by the four biobanks (LIFELINES, LLS, RS and NTR). C) Principal Variance Component Analysis (PVCA) before (red) and after (green) calibration, estimating the variance explained in the dataset by available clinical variables (e.g., sex, age, BMI, diabetes). D) Bar-plots showing the differences in men and women in the calibrated MetaboHealth in the four cohorts. E) Observed mean values of age, BMI, eGFR, hsCRP and pressure ordered following the calibrated MetaboHealth in different percentiles over the entire BIOS population. F) Observed percentage of alcohol consumption, current smoking ordered following the calibrated MetaboHealth in different percentiles over the entire BIOS population.

The construction of the MetaboHealth score as published by Deelen *et al.* [12] does not include a cross-cohort calibration, but instead standardizes the individual metabolic features per study prior to computation of the score (Figure S4). While this does make the MetaboHealth score more comparable across cohorts, and satisfactory for most meta-analysis purposes, it does also negate any real biological differences that may exist between studies. Conversely, when computing the MetaboHealth score on the calibrated data, i.e., after removing unwanted study differences and supplying all data as one dataset, it produced scores with interpretable differences and consistent trends across cohorts (Figure 2D-F). For instance, consistent with our expectation, the calibrated MetaboHealth scores now tend to be higher among the studies with the older individuals RS and LLS-PAROFFS (Figure S4A). In addition, it showed a more pronounced age-associated increase in men than in women, consistently over all cohorts (Figure 2D). Lastly, higher calibrated MetaboHealth percentiles correlated with increasing age, BMI, high sensitive CRP, and increasing prevalence of diabetes and alcohol usage (Figure 2E-F).

### DNA methylation-based predictors recapitulate metabolic markers previously associated with mortality

Our first objective was to determine if DNA methylation could simulate the MetaboHealth score, our metabolomics-based mortality predictor (Figure 1). To enforce the selection of consistent signal in different cohorts, we implement an output-specific pre-selection of consistent DNA methylation sites in the studies reserved for model development, NTR and LIFELINES (**methods**). This Epigenome Wide Association (EWAS) yielded 17,705 CpG sites showing a consistent univariate association with MetaboHealth, both in direction of association and nominal significance (p-value<0.05). Pre-selected sites were then used as input for the ElasticNET regression model predicting the MetaboHealth values (Method). The resulting model, indicated as “ *DNAm-MetaboHealth”* comprised ∼1000 sites and showed good accuracy in the 5-Fold Cross Validation test sets (5-FCV) (median r∼0.52, RMSE∼0.43), which was slightly lower, but stable, in the replication sets (LLS-PAROFF: R∼0.34, RMSE∼0.38; RS:R∼0.33, RMSE∼0.5) (Figure 3).

**Figure 3:**
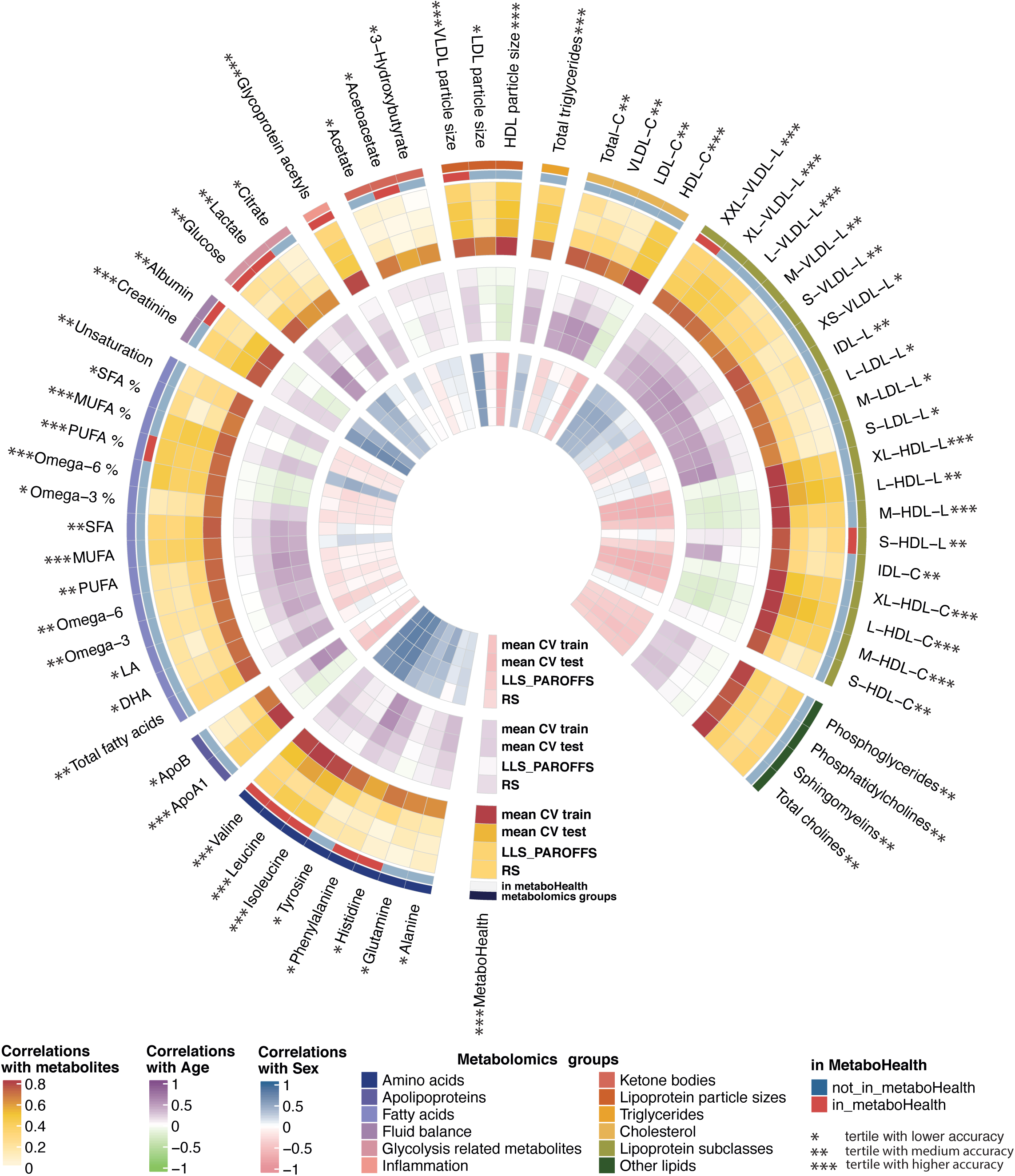
DNAm metabolites accuracies. Circular heatmap representing the accuracies of the DNAm-based models for 64 ^1^H-NMR metabolic features by Nightingale Health and MetaboHealth. The outer ring shows the correlation between measured and DNAm-based metabolomics features, while correlation between DNAm surrogates with age and sex are shown in the middle and inner ring, respectively. Mean CV states for mean cross validation results in the cohorts LL and NTR together in the training (train) and test (test) sets, while results in the left-out set are indicated with RS and LLS. Moreover, the metabolomics features are annotated for their metabolomics group type (e.g., amino acids, fatty acids etc.) and if they were or were not included in MetaboHealth. Finally, we indicated with asterisks the tertiles of mean accuracies over the test sets.

In parallel, we built distinct predictors for 64 metabolic features from Nightingale Health Plc, following the same training design as for DNAm MetaboHealth (Figure 1A). The resulting DNAm-based surrogates for the metabolomic features showed diverse mean accuracies over the different test sets (5-FCV, LLS-PAROFFS and RS), with 23 models being accurate (mean R across test sets>0.35), 20 mildly accurate (0.2>mean R across test sets <=0.35), and 21 low accuracy models (mean R across test sets <0.2) (Figure 3 and S5). In the latter group we find 5 out of 8 amino acids, several LDL-related variables, all the ketone bodies and all the glycolysis related markers. The middle group is enriched with IDL related markers, 6 out of 14 fatty acids, and 2 out of 3 glycolysis related metabolites. The accurate group of DNAm-metabolomic features included, 8 out of 10 HDL-related markers, 4 out of 8 VLDL-related molecules, glycoprotein acetyls, creatinine, 3 out of 8 amino acids, and several fluid balance markers (e.g., MUFA%, Omega6%). Higher accuracies are often accompanied by a higher correlation with age (e.g. *DNAm Leucine* and *Isoleucine*) or sex (e.g. *DNAm Creatinine*) (Figure 3, inner circles), similar to what is observed for the GrimAge DNAm based components [11]. Notably, only 7 of the most accurate surrogate markers were part of the 14 original metabolomic features composing the MetaboHealth score. Nevertheless, for 18 of the 23 most accurate surrogate markers, it was previously shown that the respective metabolic features significantly associated with mortality [12].

### DNAm metabolomics surrogates confer a unique and relevant signal

To foster the concept that DNA-methylation measurements might potentially serve as a platform to integrate biomarker signals captured from various data sources, we conducted two types of experiments. First, we ensured that the signals conveyed by our novel DNAm surrogates of metabolomic features, constitute mutually independent markers, and not a multitude of highly similar signals (Figure S6A). Then, comparing the correlations between our surrogates with the original metabolomic features (Figure S6A, upper triangle), we observed a structure remarkably congruent with the correlation structure observed between the original metabolites (Figure S6A, lower triangle), albeit overall at slightly lower magnitude. This indicates that, apart from the correlation structure between the original markers, no systematic high inter correlations are observed, which thus suggests that the information in DNA methylation measurements are sufficiently rich to reconstitute many closely related biomarker signals, without introducing artificial interdependency.

Secondly, we explored to what extent our novel DNAm surrogates of metabolomic features constitute novel signal compared to previously constructed DNA-methylation estimates. Overall, low correlation (max|R|∼0.4) are observed between our DNAm-based metabolomics surrogates and DNAm-based multivariate clocks (Horvath, Hannum, PhenoAge, and GrimAge) (Figure 4). Furthermore, correlations with other pre-trained DNAm-based surrogate molecular markers (GrimAge components and the 109 protein EpiScores) are generally modest, with a few notable exceptions. Particularly, the GrimAge surrogates DNAm-leptin and DNAm-adm, and 4 Episcores (2771.35 [Gene: IGFBP1], 4929.55 [Gene: SHBG], 3505.6 [Gene: LTα], CD6), present a relatively high positive correlation with HDL related surrogate markers and a relatively high negative correlation with the amino acids (DNAm-Leucine, DNAm-Isoleucine, and DNAm-Valine). We observe the inverse pattern for the GrimAge surrogate DNAm-PAI-1, and 4 other EpiScores (4930.21 [Gene: STC1], 2516.57 [Gene: CCL21], 3343.1 [Gene: ACY1], 3470.1 [Gene: SELE]) which also show a high correlation with VLDL surrogate markers. Notably, these correlations might suggest a link between the metabolome and protein markers related to immune signaling (LTα, CD6, CCL21, SELE), energy balance and metabolism-related hormones (IGFBP1, SHGB, ADM, leptin, and STC1), and atherosclerosis/thrombosis inhibitor (PAI1). Nonetheless, the majority of the markers exhibit limited correlations with their predecessors (Figure 4), implying the presence of previously unexplored information in DNA methylation patterns.

**Figure 4:**
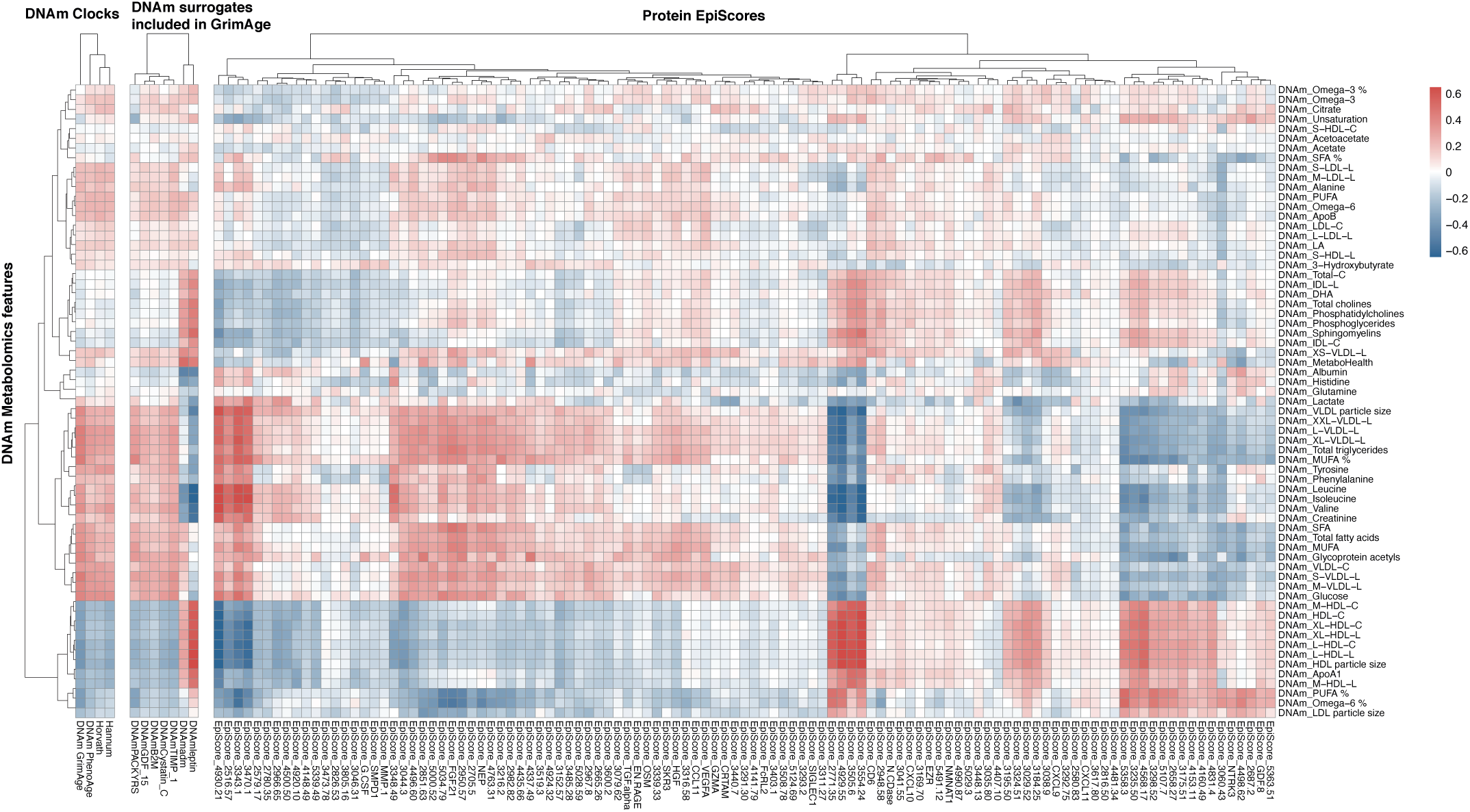
Correlations with pre-trained DNAm scores. Correlations between our DNAm metabolic features and previously trained clocks, the DNAm surrogates included in GrimAge and the 109 DNAm-based surrogates for proteins (EpiScores) by Gall et al.

### Metabolomic surrogates improve the mortality predictions of GrimAge

Next, we evaluated the DNAm-metabolomics features for their predictive value for all-cause mortality in the Rotterdam Study (RS). For this purpose, we utilized a total of 1544 samples from this cohort (mean age at baseline of 64 years, 251 deceased, and a median follow-up of 11 years; Figure 1), by incorporating an additional 863 samples with available Illumina 450k and mortality information, but not ^1^H-NMR metabolomics. In accordance with previous studies [11,14,15], we assess univariate Cox proportional hazard models (in years of follow up) adjusted for relevant covariates, specifically sex, together with age, BMI and cell counts levels at blood sampling (Figure 5A and S7A).

**Figure 5:**
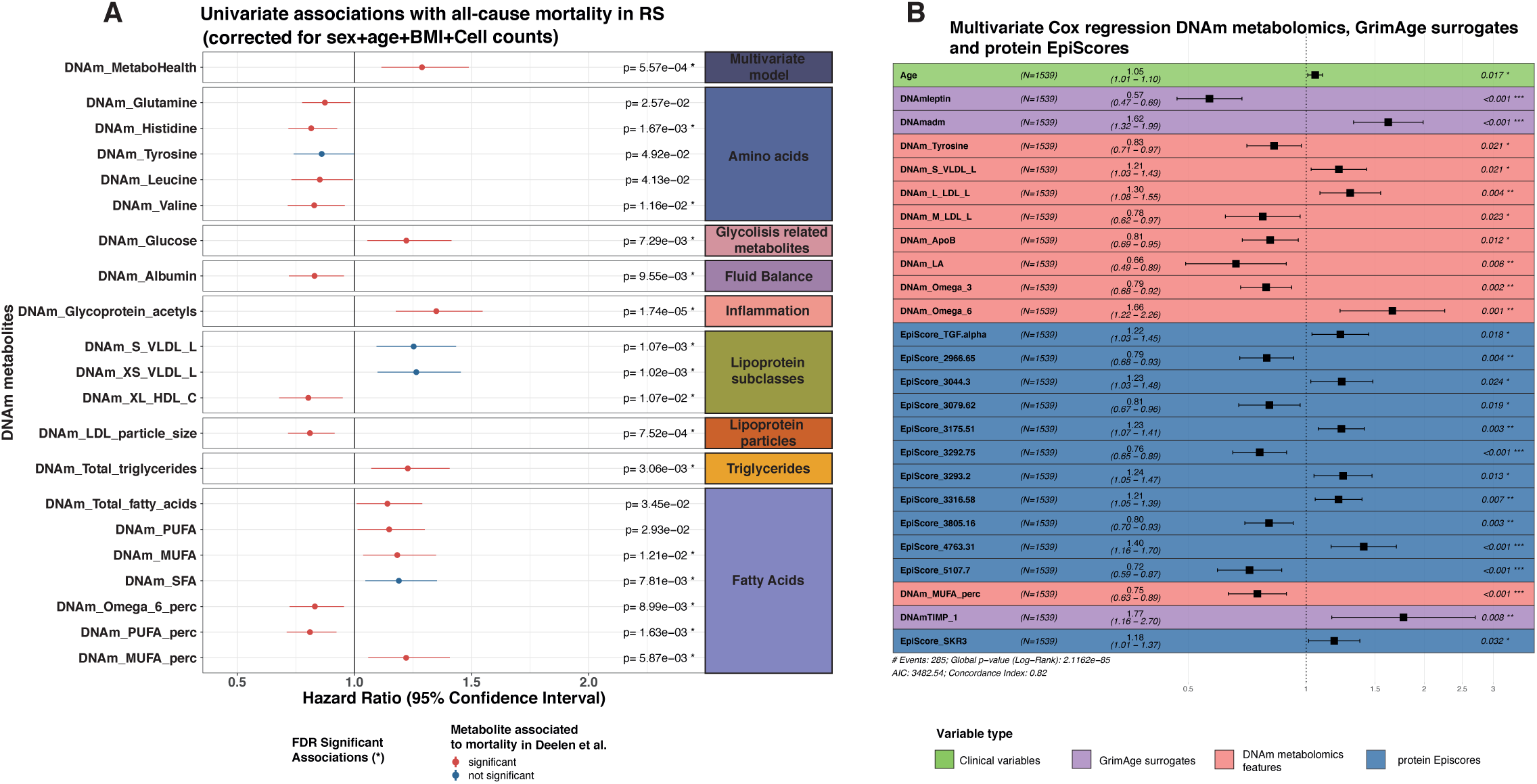
Associations with Time to Death. A) Significant univariate associations of the DNAm metabolomics features with time to all-cause mortality in RS (N = 1542 with 285 reported deaths). The associations are grouped based on the metabolomics groups colored by the significant associations or the metabolites with mortality in Deelen et al. The asterisks (*) separates nominal significant DNAm metabolomics features from the FDR significant ones. B) Stepwise Cox regression predicting of time to all-cause mortality optimized in RS, composed combining age, 3 DNAm surrogates included in GrimAge, and 9 DNAm metabolic models and 12 protein EpiScores.

First, we evaluated our *DNAm-MetaboHealth* predictor, which showed a significant association with all-cause mortality (HR=1.29, p=5.57×10^-04^) (Figure 5A), in line with the original metabolomics-based MetaboHealth score (Figure S8C). Next, we evaluated the individual DNAm metabolomics features and observed significant associations with mortality for 15 out of the 64 surrogate metabolites, of which 6 (out of 14) metabolomics features were included in the original MetaboHealth score. In addition, our estimated effects were overall consistent with those found by the study performed by Deelen *et al.* which employed a considerably larger dataset of 44,168 individuals (Supplemental S7C), with our most significant findings being amongst their strongest effects. We observed an increased risk for higher estimates of 8 DNAm-based features, with the strongest being *DNAm-Glucose* (HR=1.22, p=7.29×10^-03^), *DNAm-Glycoprotein Acetyls* (HR=1.35, p=1.74×10^-05^). Conversely, we observed protective effects for 7 features, such as *DNAm-PUFA%* (HR=0.81, p=1.63×10^-03^), *DNAm-Histidine* (HR=0.82, p=1.67×10^-03^), *DNAm-Valine* (HR=0.83, p=1.6×10^-02^), and *DNAm-Albumin* (HR=0.83, p=9.5×10^-03^). In addition, 5 nominal significant additional metabolites showed discordant mortality associations between sexes. Explicitly, for males we observed mortality associations with DNAm-Glutamine, whereas DNAm-Tyrosine, DNAm-Leucine, DNAm-Total Fatty acids, and DNAm-PUFA associated with mortality in women (Figure S7B).

Almost all the pre-trained DNAm clocks that we considered (Hannum, PhenoAge, GrimAge and bAge) and some of their intermediate surrogates exert mortality associations in RS (Figure S9A-C). Next, we attempted to refine the current standard for biological age estimation, specifically GrimAge (CI=0.79, p=4.6×10^-77^), by training a multivariate all-cause mortality predictor including our novel DNAm metabolomics features. As a first exploration, we trained a Cox regression model with age at blood sampling, sex, DNAm-GrimAge and DNAm-MetaboHealth, which only showed minor, but significant, improvements in the C-index (CI=0.8, p=4.6×10^-77^) (Figure S10 B). As a second exploration, we performed a stepwise (backward/forward) Cox regression model to identify a minimal set of features including age, sex, our 64 DNAm-metabolomic features and DNAm-GrimAge (CI=0.81, p=1.7×10^-83^) (Figure S10D). Nonetheless, the best performing model was obtained when including age, sex, 3 out of 8 GrimAge components (predicting Leptin and ADM and TIMP_1), 12 of the 109 protein EpiScores, and 9 out of 64 DNAm metabolites (CI=0.82, p=1×10^-85^) (Figure 5B). The selected DNAm metabolic features included *DNAm-Tyrosine*, *DNAm-S-VLDL-L*, *DNAm-S-LDL-L*, *DNAm-M-LDL-L*, *DNAm-APOB*, *DNAm-LA*, *DNAm-omega3* and *DNAm-omega6, and DNAm-MUFA*. In any case, all the newly introduced scores exhibited a significantly improved C-index and a higher AUC at 5 and 10 years compared to the GrimAge (Figure S10E-H). Overall, this indicates that DNAm-surrogates from different origin, phenotypic, proteomic, or metabolomic, might confer mutually independent information for mortality prediction.

### DNAm metabolomics models introduce relevant CpG selections

After establishing the value of our novel DNAm metabolomics features in predicting mortality, we explored the nature of the signal included in our models by investigating the CpG sites picked by the ElasticNET regression, which can shrink contributions of unnecessary features to zero. Predictors selected a median of ∼750 CpG-sites, with a minimum of 234 CpG-sites for *DNAm-Acetoacetate* and a maximum of 1,569 for *DNAm-ApoA1* (Figure 6B). A total of 22,145 probes were included in at least 1 model. Comparison of the genomic positions of the selected CpG-sites with the rest of the 450k array highlighted an underrepresentation of probes positioned in CpG Islands, and a preferential selection for CpG shelves and shores, known to be more dynamic areas (Figure 6A) [19,20]. Noteworthy is the higher tendency to select CpGs co-locating with enhancers, cis-acting short regions of DNA that control the temporal and cell-specific activation of gene expression (Figure 6A) [21].

**Figure 6:**
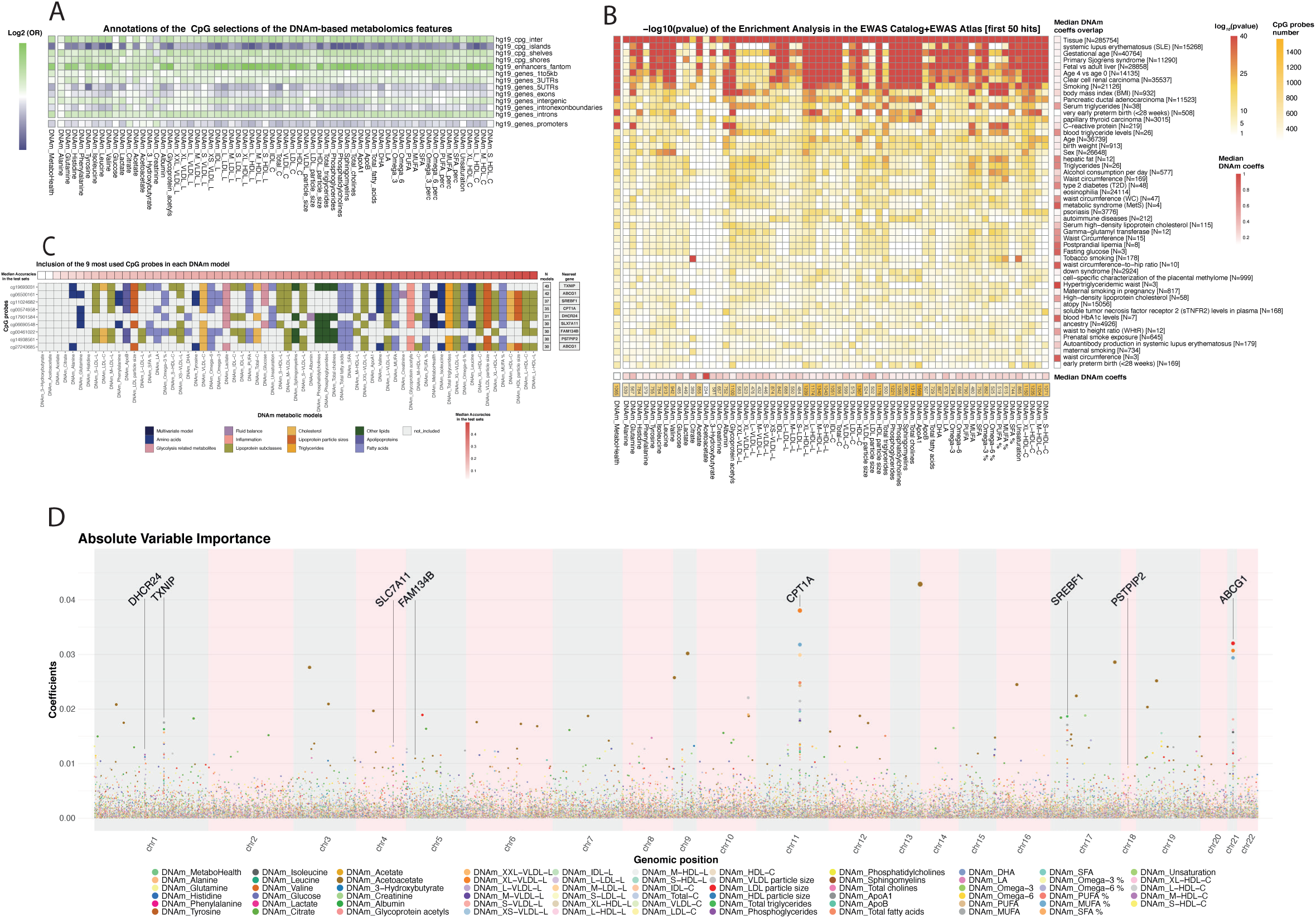
CpG selections of the ElasticNET models. A) Log2 Odds ratio indicating the enriched in annotations of the CpG by our ElasticNET models. B) The central heatmap reports the log 10 P-values of the enrichments CpG sites selected by our models (rows) and the 50 most significant traits in the EWAS Catalog and Atlas enriched (rows). Bottom: the median coefficients in each DNAm model, and the number of CpGs per model. Right: the median coefficients given by our DNAm model to the overlapping CpGs with each trait. C) The nine most used probes (rows) over the 65 ElasticNET models (columns), colored by metabolic groups. Top: The models were ordered by the mean accuracy over the test sets (CV, LLS, and RS). Right: The number of models which include each CpG and their nearest genes. D) Manhattan plot-like figure indicating the Variable importance of the single CpG probes in the DNAm metabolic models.

Functional enrichment analyses using the most proximal genes to the selected CpG-sites highlighted pathways associated to “developmental processes”, “cell differentiation”, and “regulation of metabolic processes” from Biological Processes in Gene Ontology (Figure S11C). Concomitantly, enrichment analyses of phenotypic annotations in the EWAS Catalog and EWAS Atlas (Figure 6B), indicated that the CpG-sites are known to be largely related to peripheral tissue differentiation [22], fetal brain development [23] and gestational age [24]. Nonetheless, the CpG sites with the highest median coefficients across all our models were the ones annotated for metabolite-related traits, such as “Triglycerides”, and “Fasting Glucose” (Figures 6B, S11A-B). In total, 203 traits exhibited a significant enrichment for the CpG selections made by our models. Notably we find also highly significant associations with “Ageing”, and “all-cause mortality”, indicating that we do identify CpGs related to age-related processes.

Despite their interesting overarching signal, the DNAm-metabolic models show little overlap with each other in their CpG selections, with the majority showing overlaps well below 15%, apart for a few exceptions of highly correlated metabolites (e.g., 83% between *DNAm-Total_cholines* and *DNAm-phosphoglycerides*; Figures S6 and S12). Nonetheless, a handful of CpG probes were chosen in more than 30 models with largely consistent coefficient signs (Figure 6C). Interestingly, while some of these 9 features have a higher importance weight on the DNAm metabolomics models (e.g., cg00574958, or cg06500161), others only exert a more minor influence (e.g., cg14938561, cg00461022). The 9 CpG sites with higher importance weight don’t favor one specific metabolic group but seems to be relevant to many metabolic markers (Figure 6D). Not surprisingly, also the nearest genes to these 9 probes are noteworthy. For instance, TXNIP, which includes cg19693031 (chosen in 43 DNAm-metabolomics models), was previously associated to hyperglycemia and insulin resistance, and ABCG1, nearby cg06500161 (in 42 DNAm-metabolomics models) was associated to plasma lipid levels and stroke (Figure 6D).

## Discussion

A comprehensive quantification of biological ageing, as a way to assess the overall, holistic health status and disease susceptibility of individuals [14], would constitute a major advance for healthcare and preventive research. A diversity of molecular markers has been proposed as indicators of biological age relating to health- and lifespan. Here we integrated well-established DNA methylation-based and ^1^H-NMR metabolomics resources for biological age prediction with mortality as a primary endpoint. To our knowledge, the potential synergistic effects arising from combining these two molecular sources remained thus far largely unexplored, and we believe that a collection of models predicting metabolomics features may be relevant within the rapidly growing repertoire of DNA methylation-based estimates [10,11,15]. A structured training and evaluation design aided us to demonstrate the robustness of our features. We highlighted the distinct signal expressed by our novel models and their feature selection. Finally, we explored the use of our novel DNAm-based surrogates of metabolomics features in combination with previously trained DNAm-based surrogates (e.g., Grimage constituents) suggesting that these confer complementary information.

We applied ElasticNET regression models to the data of four large population cohorts to derive DNAm-based surrogates for a previously derived multi-analyte score indicating mortality (MetaboHealth), and for 64 individual metabolomics features. The direct estimation of metabolomics-based mortality by constructing a DNAm surrogate for the MetaboHealth score showed promising results (mean R in test-sets=0.397). Moreover, we were able to construct DNAm surrogates for many, but not all, metabolomics features with good replication accuracies (mean R in test-sets>0.35), including health markers for HDL and VLDL metabolism, inflammation, and fluid balance. Less accurate were the DNAm surrogates for amino-acids, ketone bodies, glycolysis, and LDL-related markers (mean R in test-sets <0.2). Nevertheless, considering the limited number of available markers and the low accuracy thresholds previously used for DNAm scores (R>0.1 in test sets) [15], we continued evaluating all 65 models. This decision was further corroborated by a previous report by Stevenson *et al.* who suggested that their DNAm surrogate for CRP was a more reliable indication of chronic inflammation than its measured counterpart, even when considering the modest correlation between CRP and its surrogate [25]. Overall, our DNAm metabolomic features conveyed a signal coherent with the quantified metabolomics variables and independent from most of the previously reported DNA methylation-based clocks and molecular surrogates.

Great emphasis was given to the harmonization of metabolomic data collected across different cohorts, prior to training our DNAm-based models for individual metabolites or the MetaboHealth score. Non-biological variability that may originate from inter cohort differences in sample collection, storage, or handling could confound model training. Typically, this challenge in epidemiology is addressed by applying a z-scaling per cohort prior to conducting a meta-analysis, which in effect discards all differences, both technical and biological, between cohorts. In other words, while allowing to draw conclusions on the similarities in associations with endpoints between cohorts, this strategy does not allow for a direct comparison of the underlying molecular profiles between cohorts. To address this issue we applied a calibration technique, which we developed adapting methodologies previously applied in longitudinal studies [18]. This novel calibration technique showed its merit in harmonizing the metabolomics profiles, while preserving the natural biological heterogeneity within and between the different study populations. Importantly, this approach allowed for an evaluation of the MetaboHealth score across cohorts, showing consistent age and sex specific trends per study, and global predictive power for established clinical variables, such as hsCRP and diabetes.

Previous studies have shown advantages of pre-selecting CpGs when training ElasticNET regression models [26–29]. Following this example, we implemented a pre-selection of CpG sites showing a high variability and consistent association with the outcome of interest during the training phase of our 5-Fold Cross Validation procedure. Approximately 22,000 CpG sites were included in at least one DNAm-based models. Enrichment analyses showed that the selected CpGs are more likely to be enhancers in CpG shelves and shores and are in the proximity of genes enriched for regulation of metabolic and developmental processes, or cell differentiation. This finding resonates with a longstanding hypothesis, that the ageing methylome reflects processes underlying intricate cellular and molecular changes linked with development and differentiation [30]. Furthermore, CpG sites selected for our surrogates were also previously associated to age (e.g., Ageing, all-cause mortality), inflammatory (C-reactive proteins), or metabolically related traits (e.g., triglycerides and metabolic syndrome). Strikingly, we found a highly recurrent selection of 9 CpGs in at least 30 distinct DNAm surrogate models, suggesting that these CpGs form a fundamental link between the blood metabolome and DNA methylome. All these loci have been previously found associated with metabolic traits and processes [31], and most of these 9 CpGs and their nearest genes are considered powerful classifiers for diabetes stratification [32–34]. Remarkably, 3 of these 9 CpG probes showed significant univariate association with mortality within the Rotterdam Study (Figure S8D). This reassures over the valuable cardiometabolic content latent in our novel DNAm models.

Besides, our main intent was to evaluate the possibility to extrapolate the mortality signal from the metabolome to DNA methylation. To do so, we tested which of our surrogates might be indicative of all-cause mortality in a subset of the Rotterdam Study (1544 persons, 285 deaths). Notably, we observed a successful detection, albeit partial, of the mortality signal exerted by the metabolomics platform. We could successfully derive a DNAm-based version of MetaboHealth, which significantly associates with all-cause mortality, although it showed a lower hazard ratio than the original score [35]. This might in part be explained by the fact that only 6 of the 14 DNAm surrogates for the metabolites constituting the MetaboHealth showed associations with all-cause mortality. Overall, we observed significant associations with mortality for 15 out of 64 DNAm-based metabolites. The detected effects are consistent with the results previously reported by Deelen *et al.* in a large study using the original metabolomic features measured in 44.168 individuals. This consistency further underpins that DNAm surrogates for metabolomic features could potentially be leveraged as novel epigenetic markers of biological ageing.

To further explore this concept, we trained a multivariate model for all-cause mortality, that was allowed to select from all available DNAm surrogates using a stepwise forward/backward regression. This final model included 9 DNAm metabolomic features together with the competing covariates age, 3 GrimAge components and 12 plasma protein EpiScores. The resulting model combining DNAm surrogates from different origin showed a significantly improved mortality prediction (C-index=0.82) compared to the GrimAge score (C-index=0.79) (Figure 5 and S10). Our novel composite scores showed a substantial refinement of the AUC at 5 and 10 compared to the original GrimAge (Figure S10G-H). Overall, this suggests that a broader collection of DNAm-surrogates of independent origin, such as proteomics, phenotypes, and now also metabolomics, might confer a more comprehensive indication on epigenetic-based biological ageing.

An important limitation of the current study for leveraging mortality signals is its limited sample size, which is modest when compared to the large dataset that Deelen *et al.* employed to evaluate the mortality associations of the metabolomics features and to build a multi-analyte predictor for mortality. Despite the limited power, we found significant associations with mortality for the DNAm surrogates of the multi-analyte score MetaboHealth and 15 individual metabolic features, which were consistent with those observed by *Deelen et al*. A second limitation is the usage of a single endpoint, mortality, for evaluating the potential applications of our DNAm surrogates as novel marker for biological age. We acknowledge that ageing and its associated decline in overall health is a complex multi-factorial process, that is only partially captured by mortality risk. Previous work reported the merits of the ^1^H-NMR metabolomics in estimating several different types of endpoints [7,13,36,37], or even end-of-life related-phenotypes such as frailty [35], leading us to speculate that our novel DNAm surrogates for metabolomic features might also be instrumental for capturing these ageing endophenotypes.

In conclusion, we have demonstrated that metabolite markers previously associated with mortality could be leveraged to help extract the mortality signal captured by the DNA methylation platforms. Moreover, we showed that our novel DNAm surrogates capture mortality signal that is independent of the mortality signal captured by previous DNAm scores, such as GrimAge or its separate DNAm surrogate constituents. Overall, this does suggest that even more mortality signal could be extracted given the availability of proper novel mortality-associated biomarkers.

## Materials and Methods

### 1. Dataset description

#### Cohorts

This study was performed using DNA methylation data (DNAm, Illumina 450k array) and ^1^H-NMR metabolomics (Nightingale Health, platform version 2020) from 4 Dutch cohorts: LifeLines-Deep (LL), Leiden Longevity Study (LLS-PARTNER-OFFSPRINGS), Netherlands Twin Register (NTR) and Rotterdam Study (RS), all part of the BIOS consortium [16,17]. For the current study, the BIOS multi-omics compendium was further extended with 1145 samples from the NTR for which the entire process of array measurement to quality control and normalization was done together with the other BIOS-NTR samples [38], and 904 samples from the Rotterdam Study [35]. A thorough description of all cohorts and their ethics statement are provided in the Supplementary Materials. The datasets were realized by the Dutch part of the Biobanking and BioMolecular Resources and Research Infrastructure (BBMRI-NL). The final dataset contained 5,238 samples.

#### Metabolomics data

The metabolomics data was generated by the BMBRI-NL Metabolomics Consortium. The metabolic features were measured in EDTA plasma samples on the high-throughput proton Nuclear Magnetic Resonance (^1^H-NMR) platform made available by Nightingale Health Ltd., Helsinki, Finland (platform version 2020). This technique can quantify over 250 metabolic features, including also ratios and derived features. [39,40]

#### DNA methylation data

DNA methylation data for all four cohorts was generated by the subsection of BBMRI-NL named Biobank-based Integrative Omics Study (BIOS) Consortium. The DNAm was assessed from whole blood samples with an Illumina iScan BeadChip according to the manufacturer’s protocol: the Illumina HumanMethylation405 BeadChip (450k array). For compatibility with the following versions of the Illumina array, we only considered CpG sites which are available in the Illumina HumanMethylation450 BeadChip and the MeethylationEPIC BeadChip. We analysed the DNAm f3*values*, which range from 0 to 1, to indicate the proportion of methylated sites at a specific CpG in a sample.

#### Mortality data

We evaluated the associations of the DNAm-based features with all-cause mortality in a subsample of the Rotterdam Study (RS) comprising a total of 1544 samples, only 640 of which had also Illumina 450k and Nightingale Health metabolomics. The information on the vital status of the participants in RS was last updated on the 20 ^th^ of October 2022. The dataset comprehends 1544 samples, 285 of which are deceased. All the DNAm-based features were z-scaled within the RS.

### 2. Pre-processing

#### Quality control of the metabolomics dataset

To ensure the quality of our data, we applied standardized quality control processes, which have been described in previous publications (summarized in Figure S1) [7,41]. First, we limited our analyses to a subset of 65 features (out of 250), previously selected to be a mutually independent subset [7,12,41]. This selection includes fatty acids, routine lipid concentrations, lipoprotein subclasses and low molecular weight metabolites. A complete list of the variables can be found in the Supplementary Materials. In addition, pyruvate was excluded due to its high missingness in NTR (80%). Despite a small percentage of values under detection limit for acetoacetate (8% in NTR), and an even smaller percentage of outliers in glucose and xl_hdl_c (less than 0.15%), we decided to retain all other variables (Figure S1C-E). Samples with more than 1 outlier (2 from Lifelines and 1 from RS) were further removed. We then used nipals (from the package pcaMethods) to impute the 584 missing values, which accounted for 0.211% of the remaining values. The final dataset included 4,334 samples and 64 metabolic measures.

#### Quality control of the DNA methylation dataset

The quality control and normalization of the DNA methylation (DNAm) was performed using a workflow developed by the BIOS Consortium for each cohort and thoroughly described in DNAmArray (https://molepi.github.io/DNAmArray_workflow/). In brief, sample-level QC was performed with the R package *MethylAid* [42]. Probes were set to missing based on the number of available beads (≤2), intensity equal to zero, or the detection p value (p<0.01). Probes with more than 5% missing were excluded from all samples. The remaining missingness was imputed using impute.knn from the R package impute [43]. Functional normalization was then applied as implemented in *minfi*. Finally, we removed an ulterior set of ∼60,000 underperforming probes as suggested by Zhou et al. [44]

#### Calibration of ^1^H-NMR-metabolomics

To minimize any bias that may arise from batch effects among the four cohorts included in our study, we performed a cross-cohort calibration. We followed the assumption that similar phenotypic characteristics result in similar metabolomics profiles [18]. We used sex, age, and BMI as matching characteristics, given their well-known association with the metabolomic features in the Nightingale Health Platform [7,18,41,45,46]. We considered LIFELINES as our reference cohort, as it spanned a broad range of age, and BMI and had an equal representation amongst sexes. To further minimize the impact of sex on our results, we selected the subset of samples used for cross-cohort matching to have equal numbers of men and women. Following this strategy, we identified the following subsets of participants used for matching: 73 men and 73 women in LLS-PAROFFS; 140 men and 140 women in NTR; 37 men and 37 women in RS (Supplementary Figure S2).

Based on these matching samples between cohorts, we calculated the shift in mean and standard deviation for each metabolic feature required to transform the distribution of values observed in a cohort to match the distribution in the reference cohort. We then applied this transformation to all samples of each cohort (see **Supplementary Materials**). The final dataset was log-transformed and standard normalized (zero mean and unit standard deviation) across all samples to obtain normally distributed concentration with comparable ranges across all metabolic features.

T-distributed stochastic neighbor embedding (tSNE, R package *Rtsne*) was used to visually inspect the effect of this calibration, comparing the sample similarities before and after calibration. Moreover, K-nearest neighbor batch effect test (kBET, R package *kBET*) was applied to the matching samples of each biobank before and after calibration to quantitatively evaluate the mixing of the samples [47]. Finally, we used principal variance Component Analysis (PVCA, R package *pvca*), to determine if the calibration disrupted the sources of variability of the dataset [48].

### 3. Application of previously trained multivariate models

#### MetaboHealth

The MetaboHealth model is a mortality predictor based on Nightingale Health metabolomics concentration [12]. We applied this model both on the uncalibrated and calibrated version of the ^1^H-NMR metabolomics dataset using the R-package *MiMIR* [49].

#### Epigenetic clocks

We projected the Horvath, Hannum, DNAm PhenoAge in our data using the R package *methylclock* and the DNAm GrimAge clocks using Python scripts provided by Lu et al. [11,50]. About 1000 CpG sites needed to calculate these biological ages were missing in our cohorts, therefore we imputed them using the “datMiniAnnotation3_GOLD.csv” file, dispatched by the same authors [11].

#### EpiScores

We projected the EpiScores and bAge score using the code available from the work of Bernabeu et al. [28].

### 4. Estimation and evaluation of the epigenetic-based metabolic features

We derived prediction models for the 64 metabolomics features and the MetaboHealth score using blood methylation data. For model development and testing we used NTR, and LIFELINES, respectively the largest cohort and the calibration’s reference cohort. We employed ElasticNET regression from the R package *glmnet* to train the models.

Other studies show the benefit of pre-selecting the features before using ElasticNET regression [28,51]. For this reason, we performed Epigenome Wide Association studies (EWAS) to identify CpG sites showing linear association with each feature separately in NTR and LIFELINES (*metabolic feature ∼ CpG site*). We selected the CpG probes with a consistent association sign (positive or negative in both cohorts) and significant nominal p-value (<0.05), to avoid excluding too much information.

We used a nested 5-Fold-Cross-Validation (5-Fold CV), to evaluate the models in the external loop and using the internal loop to optimize the A parameter, which determines the final set of CpG sites included in each model. The mixing parameter alpha was fixed at 0.5, based on previous work [11,41]. The final ElasticNET models were obtained using both NTR and LIFELINES and the optimized parameters. LLS-PAROFFS and RS were used as replication datasets. Finally, we report Pearson correlations (R) and the root mean square error (RMSE) of the predicted *DNAm metabolic features* with their measured concentrations.

### 5. CpG sites characterization

To gain more insight into the biological phenomena that characterize our novel DNAm metabolomics models we evaluated their fully data-driven selection of 22,145 CpG sites.

#### EWAS enrichment analysis

We utilized the MRC-IEU EWAS Catalog [52] and the EWAS Atlas [53] to assess the previously known phenotypic annotation (traits) of the CpG sites selected by our models. Both the EWAS Catalog and EWAS Atlas are online databases that compile results from Epigenome Wide Association Study results. By merging these two resources, both downloaded on March 13 ^th^ 2023, we aimed to gather a comprehensive list of previous EWASs. We gathered only the associations conducted with Illumina 450k and accompanied with a PMID. We then excluded redundancy between the two catalogs and applied Bonferroni correction using the number of CpGs in the Illumina 450k (∼480.000). This process yielded 742635 CpG-trait associations.

Next, we employed Fisher’s exact test to assess the enrichments of each of the CpG sites selected by our DNAm models. To account for multiple testing, we used the Benjamini-Hochberg correction.

#### Annotation of the genomic position of the CpG sites

We used the R package *annotatr* to annotate the genomic features. CpG sites from the 450k array were annotated using CpG Island (CGI) centric categories. The annotations we utilized are as follows: CGI (annotated in the R package *AnnotationHub*), shores (2Kb upstream or downstream the CGI), shelves (2kb flanking the CpG shores), interCGI (the rest of the CpGs). As for the genic annotations we considered regions 1-5Kb upstream of the transcription starting site (TSS), promoters (<1Kb upstream of the TSS), 5’UTR, 3’UTR, exons, introns, boundaries between introns and exons, and intergenic regions. Additionally, we report the annotations of active enhancers determined by Anderson et al. [21]

For the enrichment analyses of the annotations described above we calculated odds ratios (OR) of the CpGs included in each model compared to the rest of the 450k array. Statistical significance was evaluated using the Fisher’s exact test. The significance of the associations was established with an FDR<0.05.

#### Gene Ontology enrichment analyses

To gain further insights into the genetic context of the set of CpG sites selected, we investigated the genes in cis, considering a maximum distance of 100kB of distance.

Next, we utilized the genes associated with each CpG selections from our models to perform a functional enrichment using Gene Ontology. We employed GOfuncR package to explore the Biological *Processes* and *Molecular Functions*. The significance of the associations was established with an FDR<0.05. This analysis resulted in 2,365 significant associations between CpG sites and genes.

### 6. Associations with mortality in the Rotterdam Study

#### Univariate mortality associations

We used Cox Proportional hazard to univariately associate our 65 DNAm metabolomics features, the pre-trained DNAm clocks (e.g., PhenoAge, GrimAge), and 109 protein EpiScores with mortality (see **Supplementary Materials**). All models were corrected for age at blood sampling and sex. Additionally, we evaluated the association with mortality of our DNAm metabolomics features when correcting for sex, age and GrimAge. All *p-values* were corrected using Benjamini Hochberg and considered significant if the FDR<0.05. We used the R-package *survival* to calculate the Cox regressions.

#### Multivariate mortality models

We then combined the DNAm features with sex and age in 4 different stepwise Cox regression models (see **Supplementary Materials**). The first base model included our novel DNAm metabolomics features. The second and third model added to the first model respectively DNAm-GrimAge and the DNAm-based components of the GrimAge model. Finally, the fourth model is based on a combination of our DNAm metabolomics features, the DNAm-based components of the GrimAge and the DNAm-based protein EpiScores.

To select the interesting DNAm surrogate, we used a stepwise (backward/forward) procedure for each Cox regression model. For each of the above-described selections, we started from a model containing the full set of variables and we removed or added an unselected metabolic surrogate at each round based on the improvement on the model calculated from the C-index, taking also into account the significance of the *p*-value of each variable included in the model.

To compare the performances of the Cox regression models we used the R package survcomp within the Rotterdam Study [54]. We compared the C-indices of the newly developed models with baseline (GrimAge) using a Student t-test as described in Haibe Kans et al. [55]. Moreover, we plotted the ROC curves at 5 and 10 years of all the models.

## Data Availability

All data produced in the present study are available upon reasonable request to BBMRI-NL and BIOS.

https://www.bbmri.nl/services/samples-images-data/request-portal

https://www.bbmri.nl/bios-consortium

## Data sharing

BBMRI-nl and BIOS-nl data are available upon request at https://www.bbmri.nl/services/samples-images-data. All DNAm metabolomics scores can be obtained with a script at: https://github.com/DanieleBizzarri/DNAm_metabolomics_scores.

## Acknowledgements

This work was performed within the BBMRI Metabolomics Consortium funded by: BBMRI-NL (financed by NWO 184.021.007 and 184.033.111), X-omics (NWO 184.034.019), VOILA (ZonMW 457001001) and Medical Delta (METABODELTA: Metabolomics for clinical advances in the Medical Delta). EvdA is funded by a personal grant of the Dutch Research Council (NWO;VENI:09150161810095). Acknowledgements for all contributing studies can be found in the Supplementary Material-BIOS Consortium. Additional NTR samples were funded by the European Research Council (ERC-230374) project Genetics of Mental Illness (Boomsma).

## Contributors

EbvDA, DB, MJTR and PES conceived and wrote the manuscript. DB performed the analyses. EBvDA and MJTR verified and supervised the analyses. PES, MB, JBJvM, JvD, DIB, RP, MG, LF were involved in data acquisition of the cohort data. All authors discussed the results and contributed to the final manuscript.

