## Supplementary Figures for "^1^H-NMR metabolomics-guided DNA methylation mortality predictors"

#### **Supplementary Figure 1:** Pre-processing of the metabolomics dataset

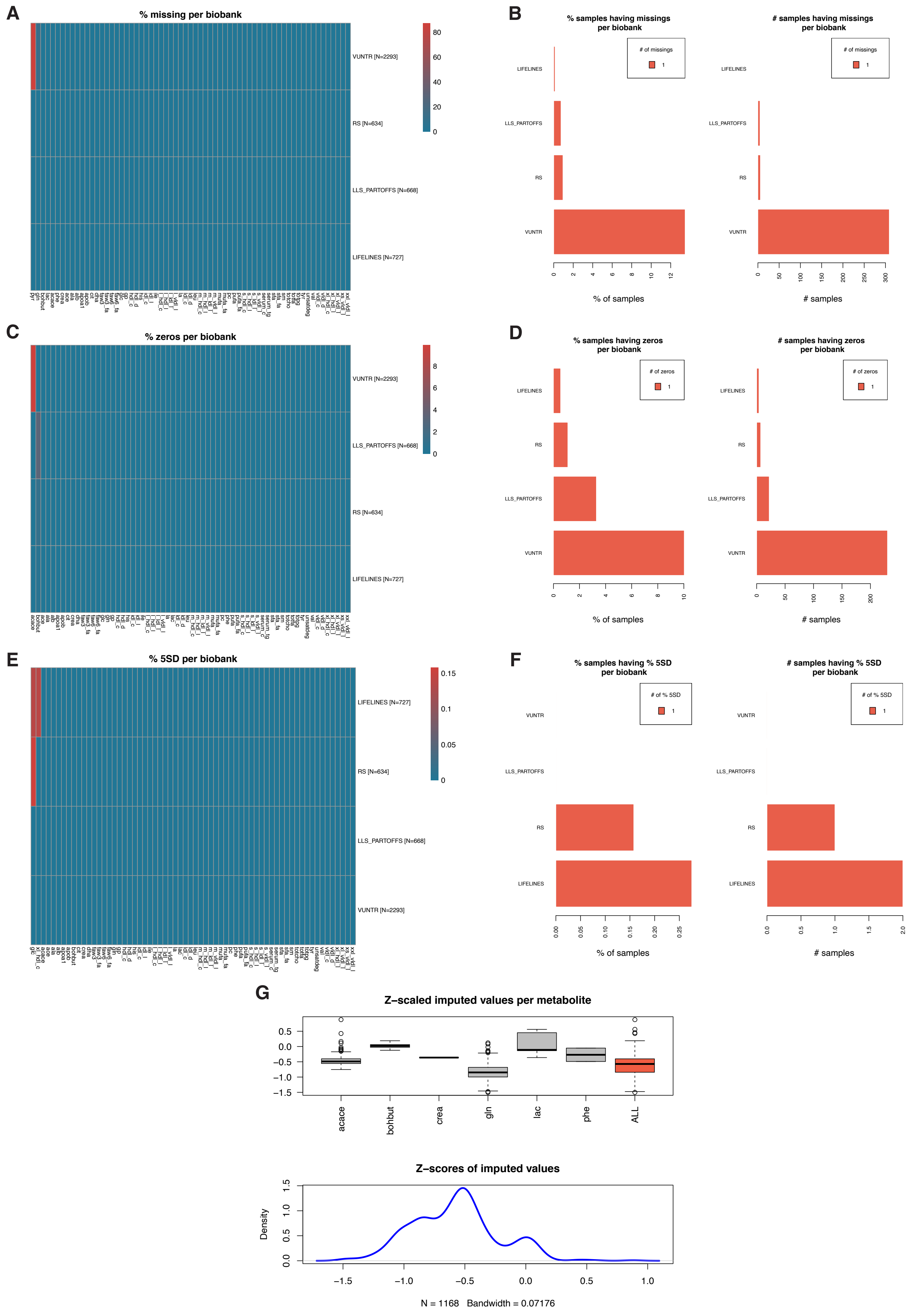

**Supplementary Information 2:** The calibration theory and the matches in LLS, LL and VUNTR

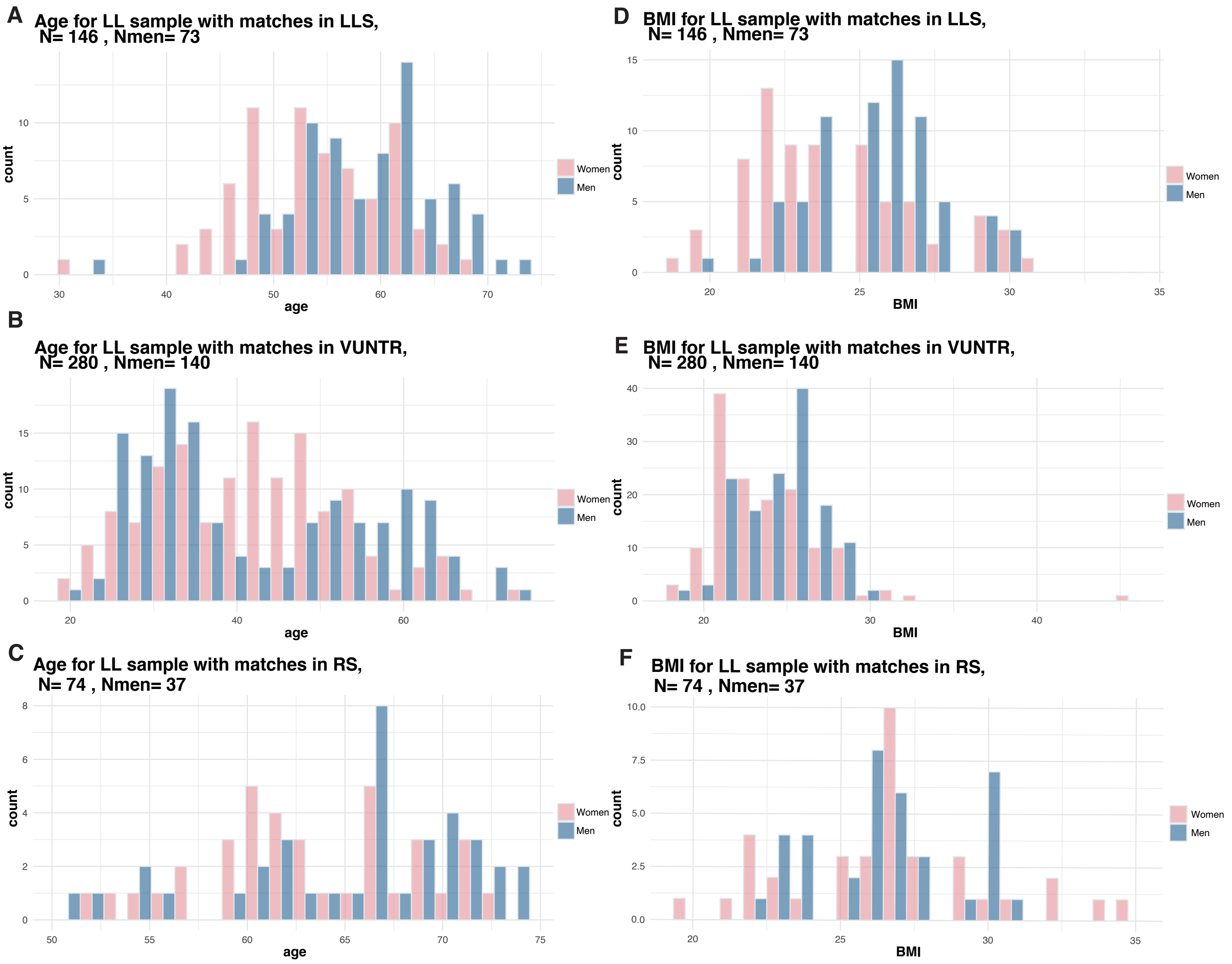

**Supplementary Information 3:** Calibration comparisons in the metabolomics

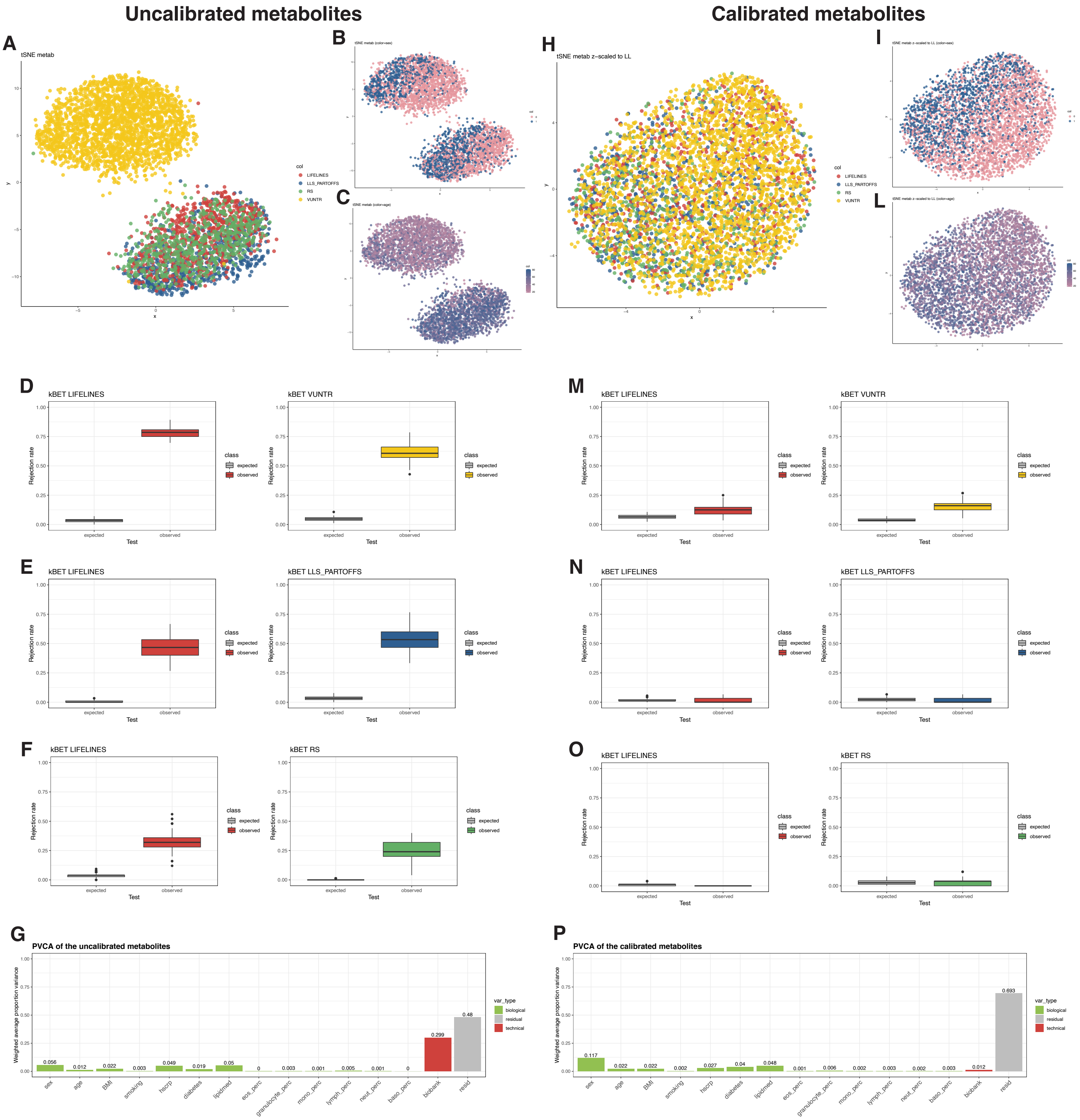

**Supplementary Figure 4:** Calibrated and uncalibrated MetaboHealth

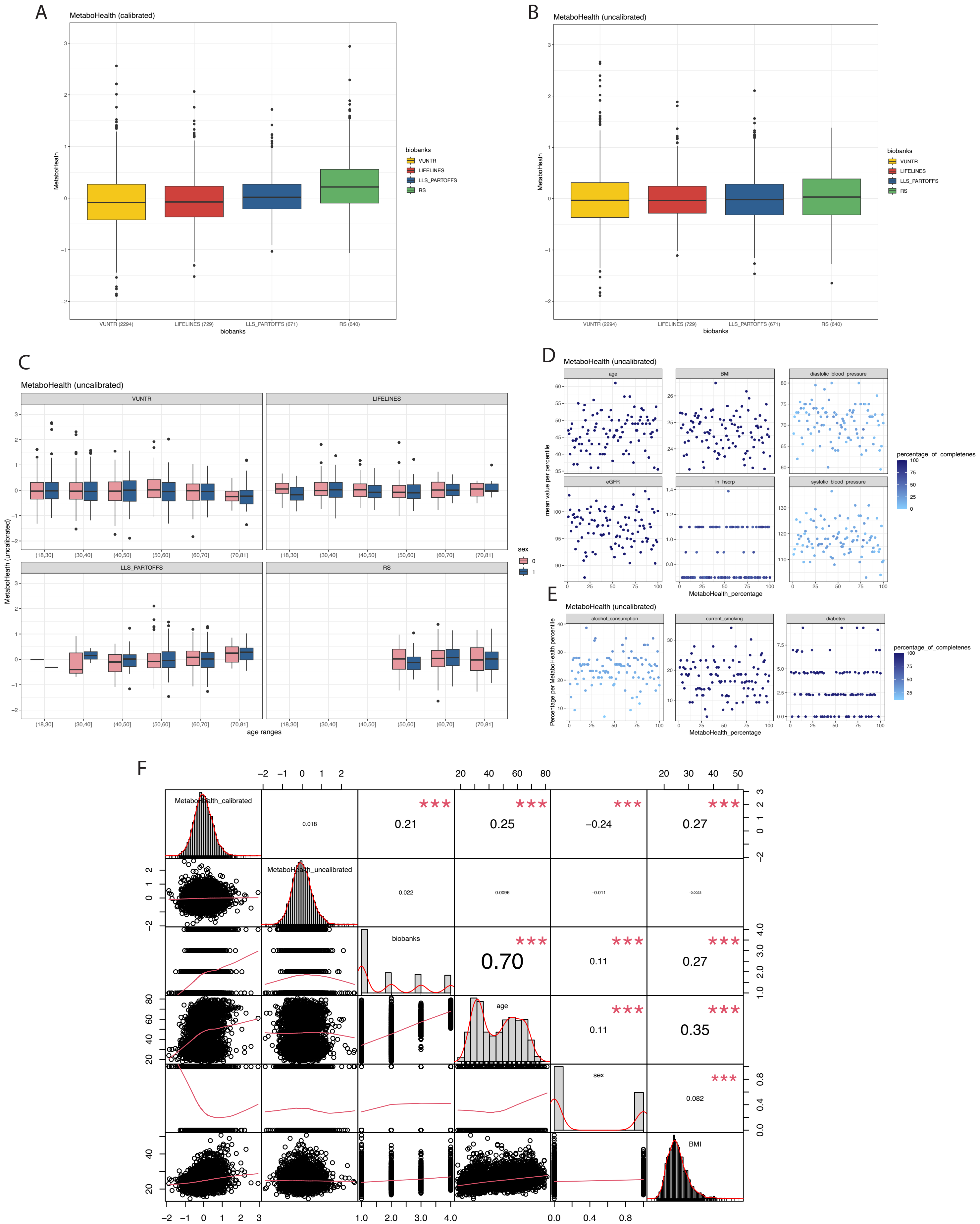

**Supplementary Figure 5:** DNAm metabolomics features accuracies divided in tertiles

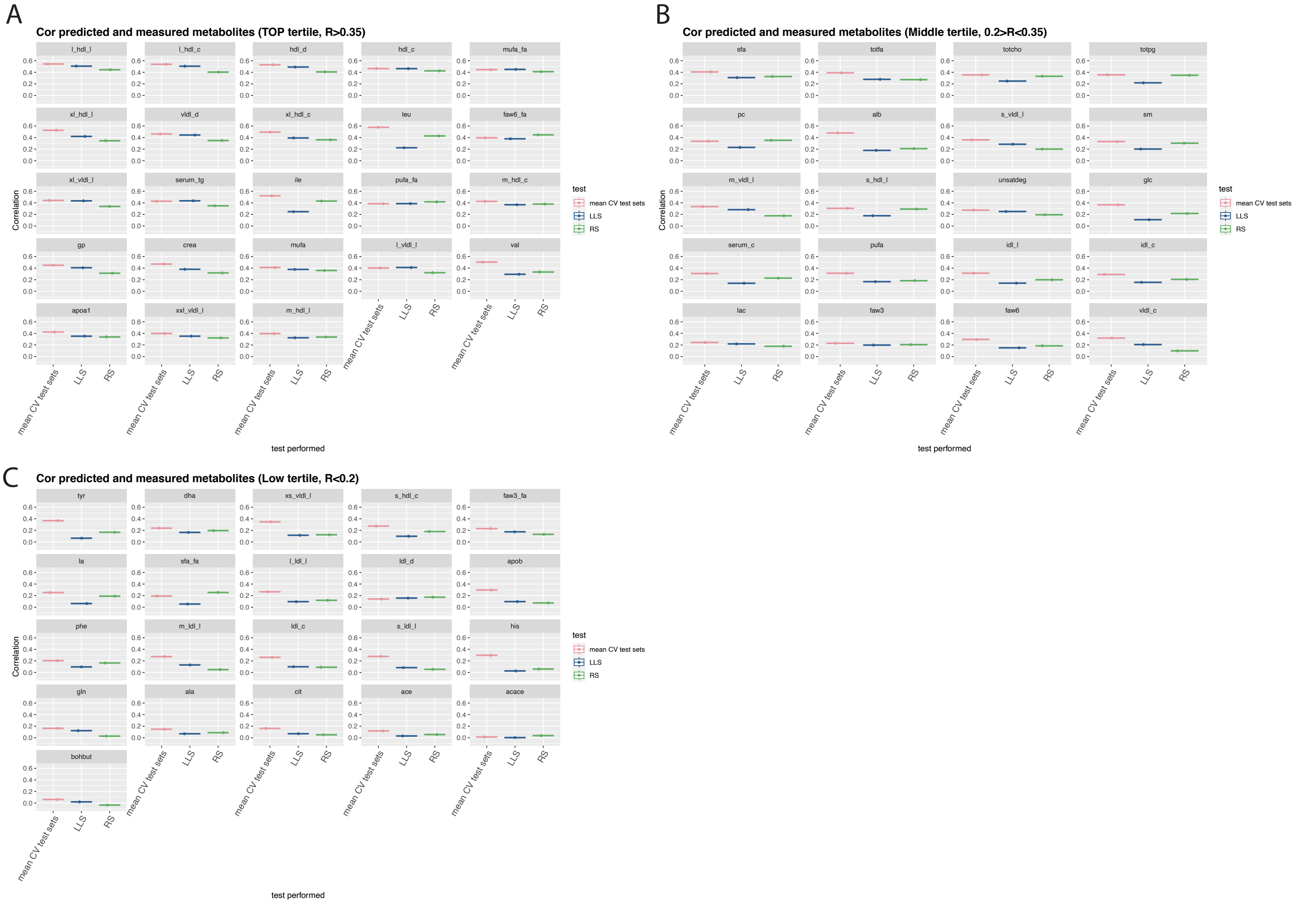

**Supplementary Figure 6:** Metabolites intercorrelations and DNAm metabolites intercorrelations

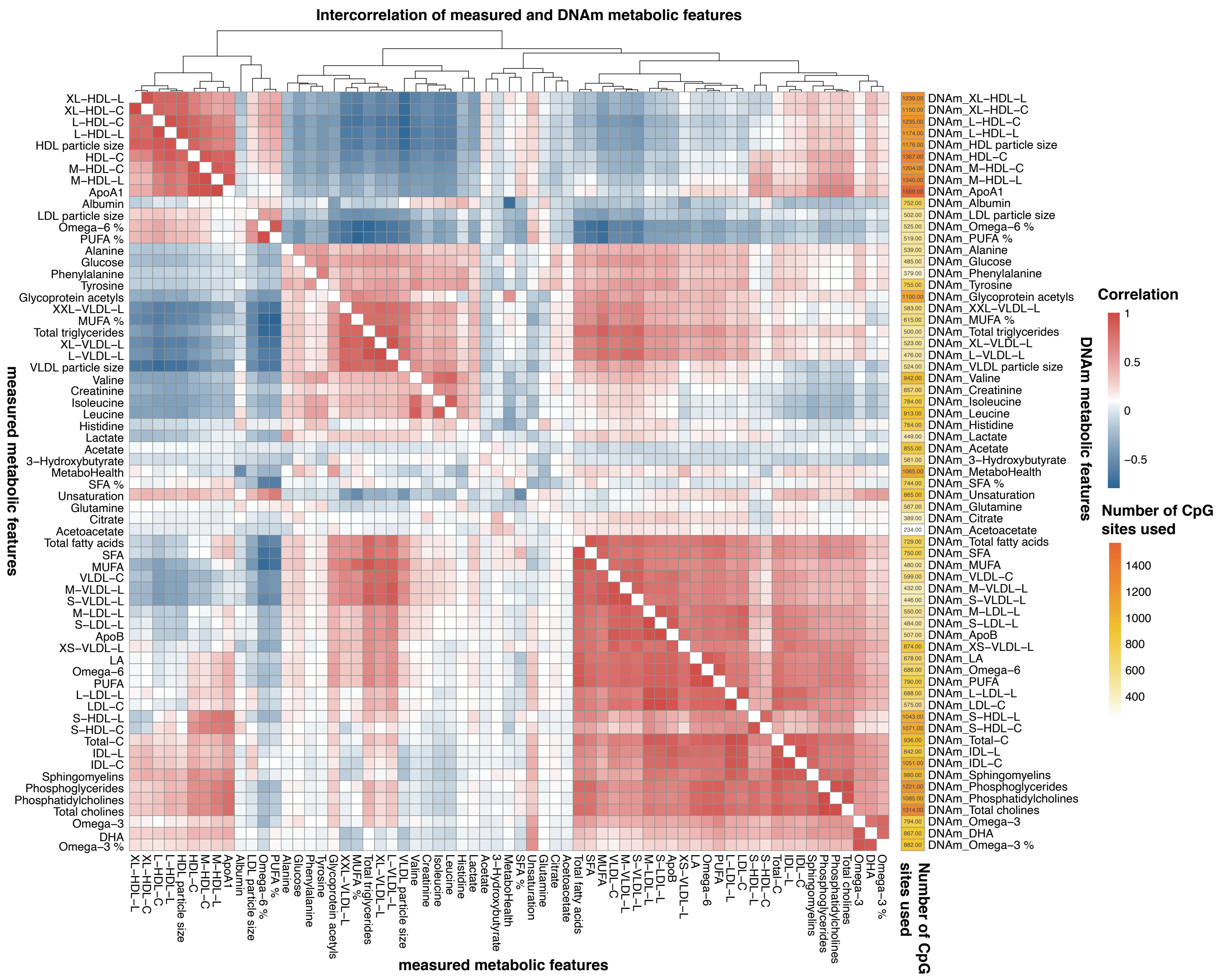

**Supplementary Figure 7:** Univariate mortality associations of the DNAm-metabolomics surrogate in RS

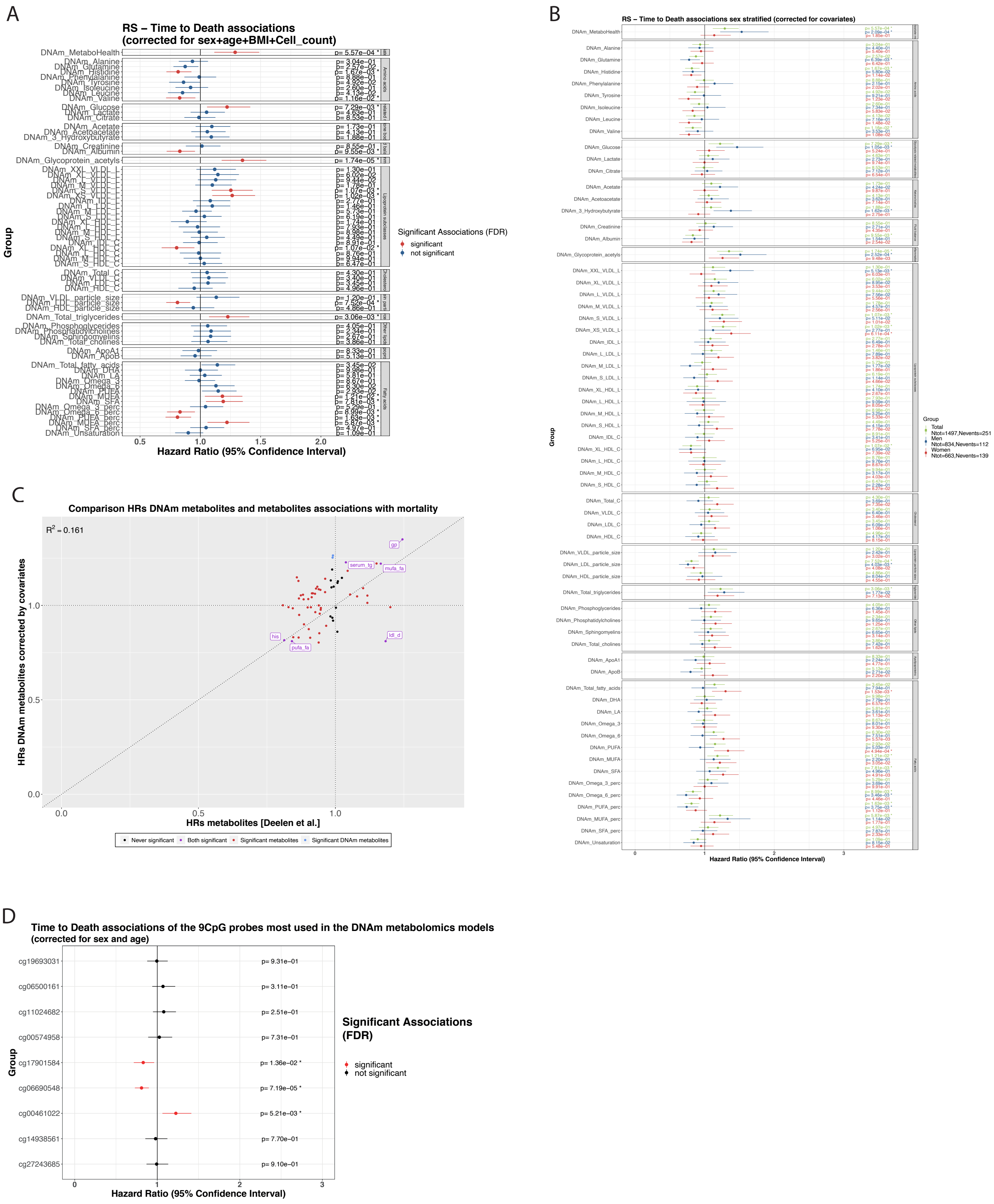

**Supplementary Figure 8:** Mortality associations of the measured metabolomics in RS

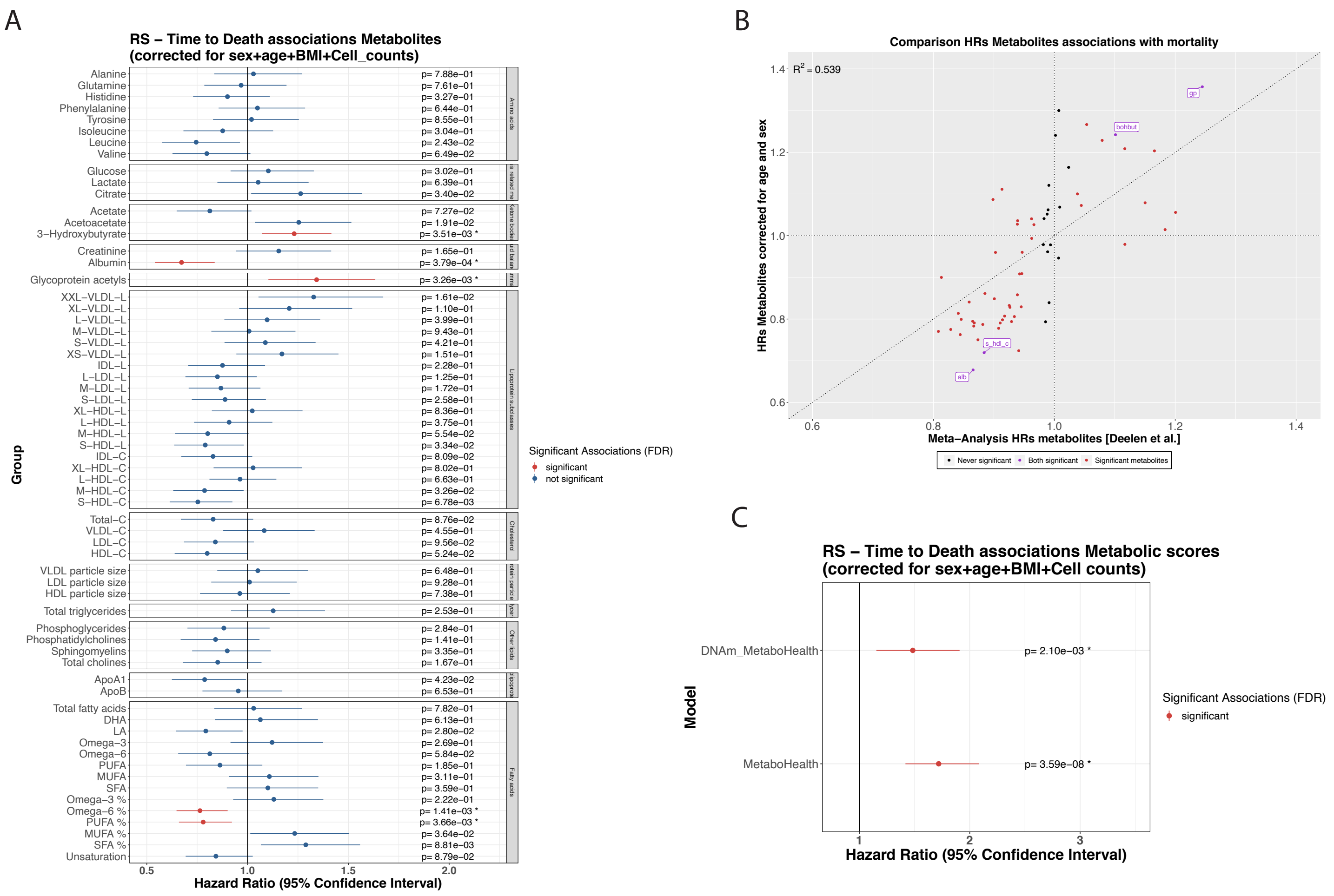

**Supplementary Figure 9:** Univariate mortality associations in RS of the pre-trained scores

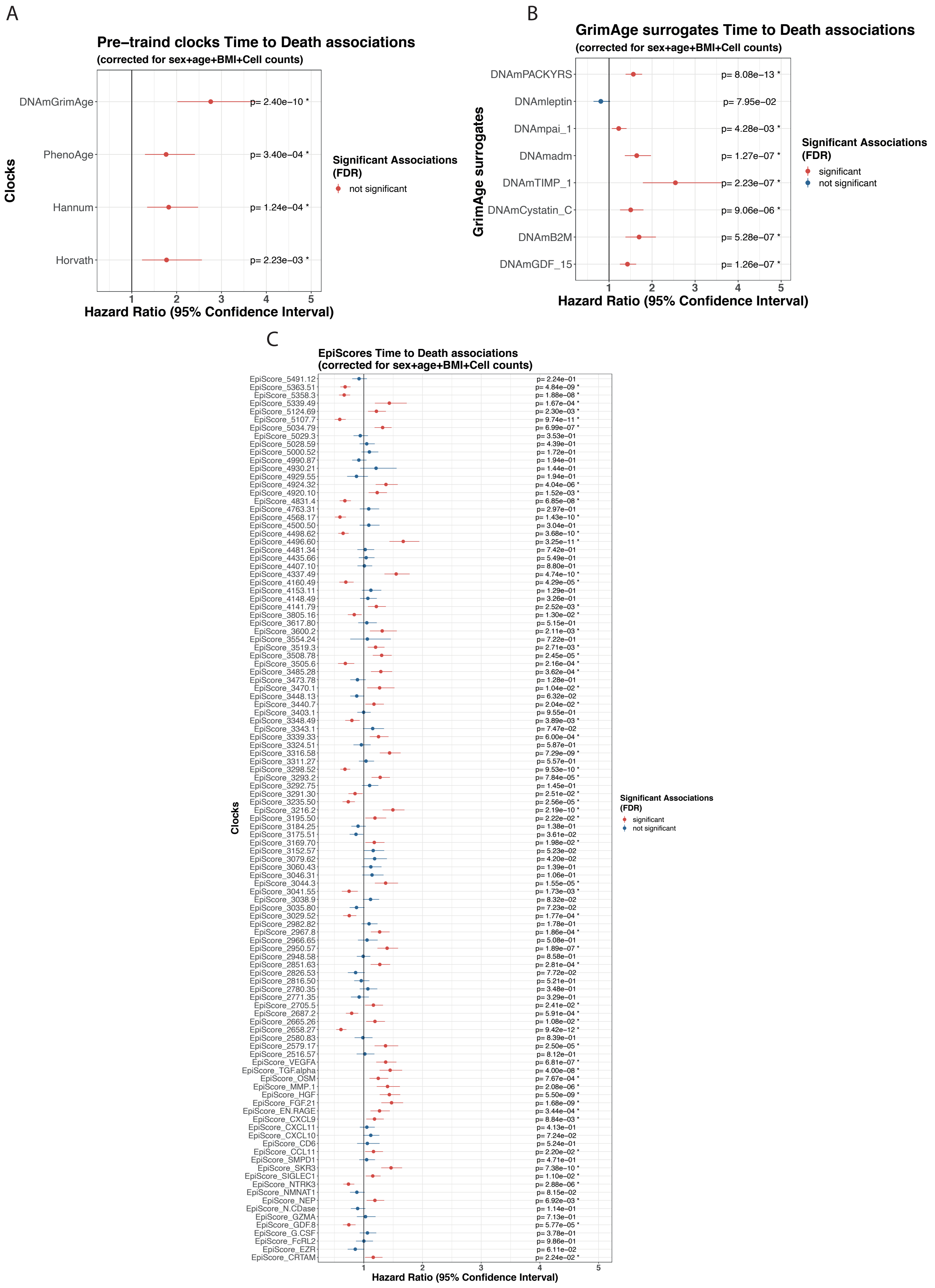

**Supplementary Figure 10:** Multivariate mortality modelsF

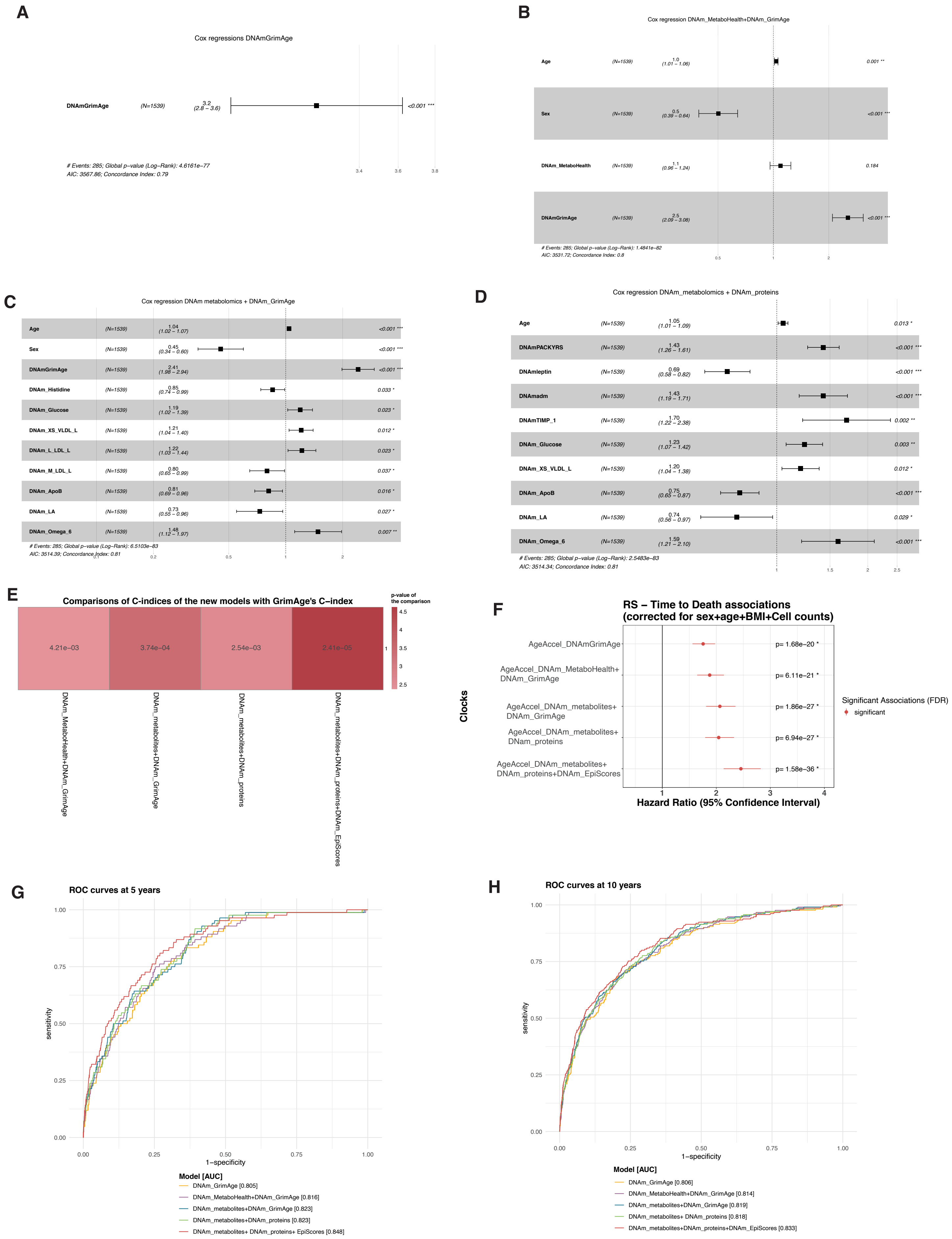

**Supplementary Figure 11:** Enrichment analyses of the CpGs selected by the DNAm models

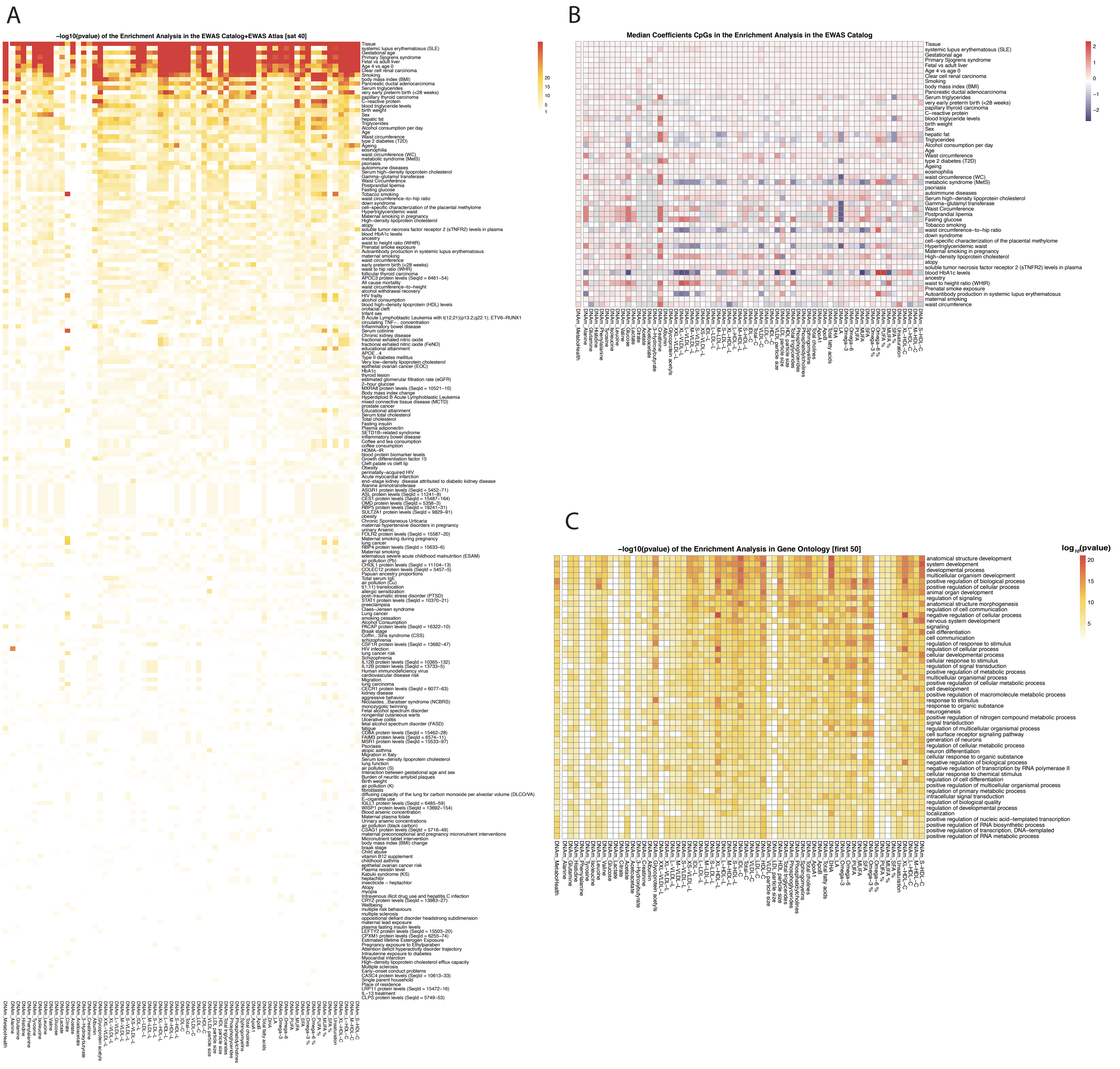

**Supplementary Figure 12:** Overlap of the selected features of DNAm metabolites

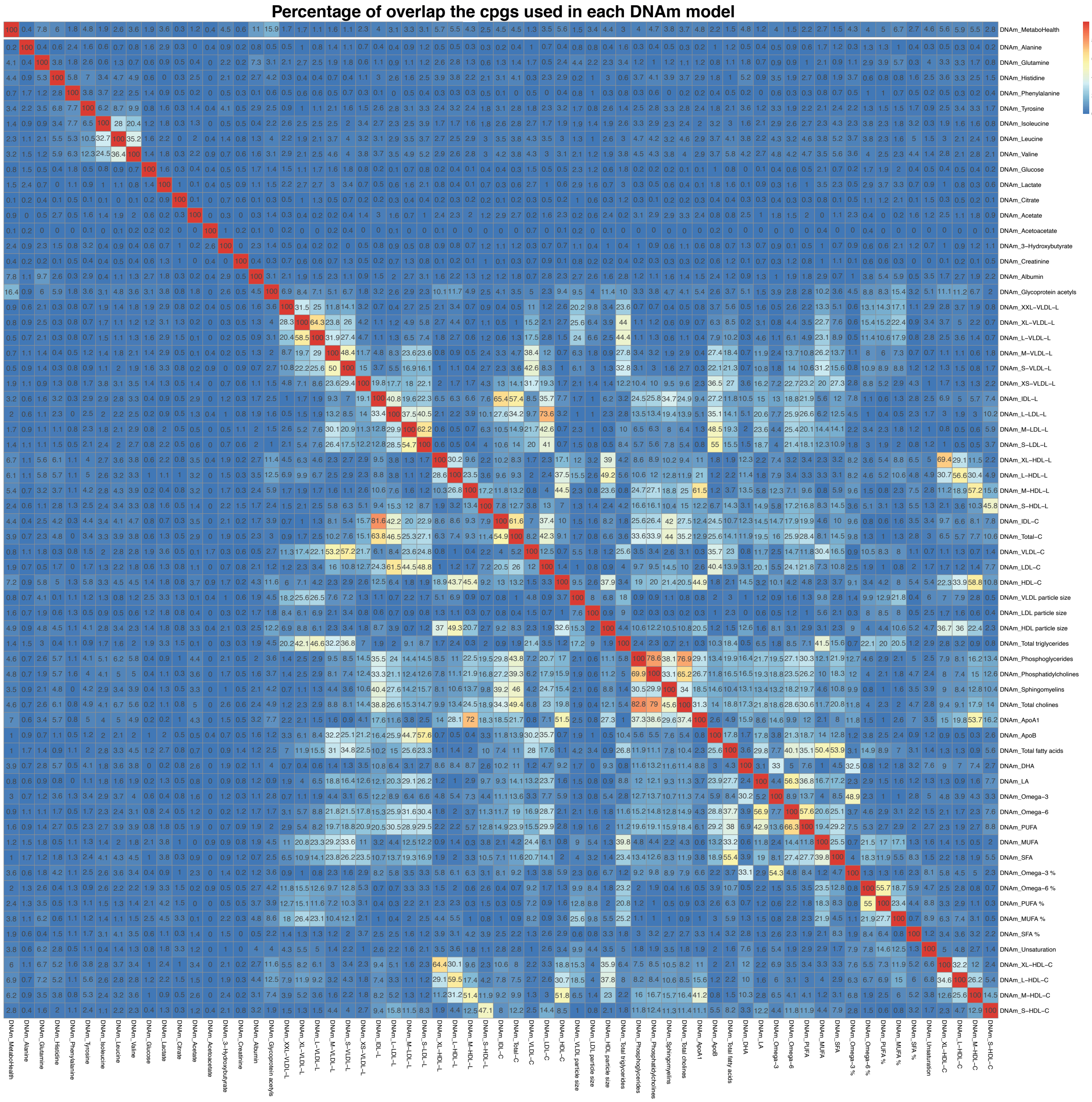

### Supplementary Figures description

**Supplementary Figure 1:** Preprocessing of the metabolomics dataset. Percentages of A) missing values. C) zeros, and E) outliers, in each 64 metabolomics features in the 4 BIOS cohorts (VUNTR, RS, LLS_PAROFFS and LIFELINES). Bar-plots representing the number of B) missing values, D) zeros, and F) outliers in the samples divided per cohort. Finally, G) indicates the Z-score distributions of the 1168 imputed missing values in the dataset.

**Supplementary Figure 2:** Distributions of the matches used to perform the calibration on the metabolomics dataset. The number of matching samples between A) B) and C) are the age distributions of the matching samples. D), E) and F) are the BMI distributions of the matching samples.

**Supplementary Figure 3:** Comparisons of the metabolomics datasets before and after the calibration. tSNE plots colored by biobanks, gender, and age before (respectively A, B, C) and after (respectively H, I, L) the calibration. kBET shows an improved mixing in the matching samples between LIFELINES and VUNTR (D before and H and after), LLS LIFELINES and LLS_PARTOFFS (E before and N after), and RS and LIFELINES (F before and O after). PVCA before G. and after P. calibration.

**Supplementary Figure 4:** Distributions of the uncalibrated MetaboHealth. Box-plot comparing the A) calibrated and B) uncalibrated MetaboHealth values in each biobank, C) Bar-plots showing the differences in men and women in the uncalibrated MetaboHealth in the 4 cohorts. D) Observed mean values of age, BMI, eGFR, hsCRP and pressure ordered following the uncalibrated MetaboHealth percentile over the entire BIOS population. E) Observed percentage of alcohol consumption, current smoking, and ordered following the uncalibrated MetaboHealth percentile over the entire BIOS population. F) Correlation chart comparing the calibrated and uncalibrated MetaboHealth, with age, sex, BMI and biobanks.

**Supplementary Figure 5:** Correlation of the DNAm metabolomics features with their quantified counterpart over the test sets (5Fold Cross-Validation test sets, LLS, and RS) divided in tertiles. Explicitly A) shows features with mean R >0.35, B) 0.2>mean R<0.35, C) mean R<0.2.

**Supplementary Figure 6:** Intercorrelations of the DNAm metabolomics features. A) Clustered intercorrelations between the DNAm metabolic features in the upper triangle and intercorrelations between the measured metabolomics features in the lower triangle. On the right there is also an heatmap indicating the number of CpGs used per model.

**Supplementary Figure 7:** Univariate mortality associations in RS. A) Complete Univariate associations of the DNAm metabolomics features with time to all-cause mortality in RS (N = 1542 with 285 reported deaths). The associations are grouped based on the metabolomics groups and colored by the significant associations or the metabolites with mortality in Deelen et al. B) Univariate mortality associations in RS corrected by sex and age of: B) the CpG probes most used in the DNAm metabolomics features; C) the pre-trained clocks; D) the pre-trained surrogates from GrimAge; E) the 109 plasma protein EpiScores.

**Supplementary Figure 8:** Mortality associations of the measured metabolomics in the overlapping 664 samples (99 deceased) in the Rotterdam Study. A) Univariates associations with mortality of the metabolomics features divided in metabolomics groups. B) Comparison of the HRs of the metabolomics features in the RS with what previously reported by Deelen et al. C) Comparison of the univariate mortality associations with MetaboHealth and DNAm_MetaboHealth.

**Supplementary Figure 9:** Univariate mortality associations in RS of the pre-trained scores. A) Univariate associations of each of the evaluated DNAm-based clocks (GrimAge, PhenoAge, Hannum and Horvath). B) The mortality associations with the DNAm surrogates included in GrimAge. C) The mortality association for each of the 109 plasma protein EpiScores.

**Supplementary Figure 10:** Multivariate mortality models built in the Rotterdam Study. A) Cox regression using GrimAge. B) Cox regression including age, sex, DNAm MetaboHealth score and GrimAge (DNAm_MetaboHealth+DNAm_GrimAge). C) Stepwise cox regression built with the DNAm metabolomics features and the GrimAge score (DNAm_metabolites+ DNAm_GrimAge). D) Stepwise cox regression built with the DNAm metabolomics features and the DNAm proteins surrogates included in GrimAge (DNAm_metabolites+ DNAm_proteins). E) P-values evaluating the significance of the improvement in the C-indices of the newly developed multivariate mortality models compared with GrimAge. F) Hazard Ratios of the clocks regressed for age, sex, BMI, and cell counts. G and H) ROC curves showing the prediction accuracy at 5- and 10-years mortality of all the clocks.

**Supplementary Figure 11:** Enrichment analysis of the CpGs selected by DNAm models. A) Log10 p-value of all the significant associations in the enrichment analysis over the selected CpG in the EWAS Catalog and Atlas. B) Median coefficients of the CpGs in the DNAm metabolomics models for the overlapping CpGs with the first 50 significantly enriched traits in the EWAS Catalog and Atlas. C) Log10 p-value of the first 50 Gene Ontology traits enriched with the nearest genes to the selected CpG sites.

**Supplementary Figure 12:** Percentage overlap of the CpG sites selected by each DNAm model.
