## Supplementary Materials for "^1^H-NMR metabolomics-guided DNA methylation mortality predictors"

### Supplementary Material of $^1$H-NMR metabolomics guided methylation mortality predictors

### Supplementary Methods

#### Harmonization of the metabolomics

We decided to calibrate the Nightingale Health metabolomics features to correct for any batch differences visible in the dataset. We started from the assumption that individuals with the same phenotypic characteristics, should have similar -omics profiles [1]. We used sex, age and BMI as matching characteristics. We considered LIFELINES our reference cohort because it spanned a broad range of age and BMI. To further minimize the impact of gender on our results, we optimized each matching subset to have the same number of men and women. We individuated the following subsets of participants: 73 men and 73 women in LLS-PAROFFS; 140 men and 140 women in VUNTR; 37 men and 37 women in RS (Figure SM1).

For each matching subset e calculated mean and standard deviation of each metabolomics feature separately. Then, using their mean and standard deviations calculated above, we applied the following calculation to the entire cohort of LLS, VUNTR and RS.

$$z_{ij} = \frac{x_{ij} - \overline{x_J}}{s_{x_j}} s_{y_j} + \overline{y_J}$$

where, $z_{ij}$ is the value of a feature for the i$^{th}$ sample in the j$^{th}$ feature after calibration, $x_{ij}$ its corresponding raw value, $\overline{x_J}$ and $s_{x_J}$the mean and standard deviation of the j$^{th}$ feature in the matching samples of the cohort to calibrate, $\overline{y_J}$ and $s_{y_j}$ the mean and standard deviation of the j$^{th}$ feature in the corresponding matching samples of the LifeLines cohort. In this way we obtained a z-scaling toward the mean and the standard deviation of LifeLines for all the rest of the cohorts (Figure SM1). The final dataset was log-transformed and z-scaled over all samples to obtain normally distributed concentration with comparable ranges across all features.

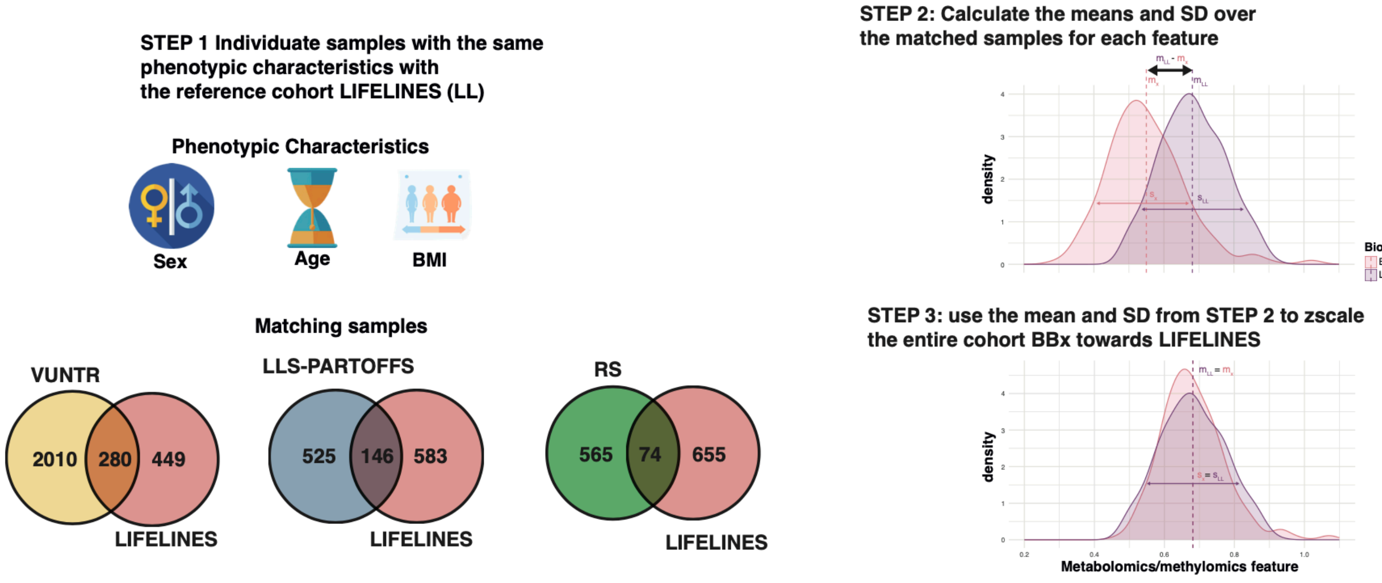

*Figure SM1: Visualization of the Calibration procedure.*

### Distributions Before calibrations

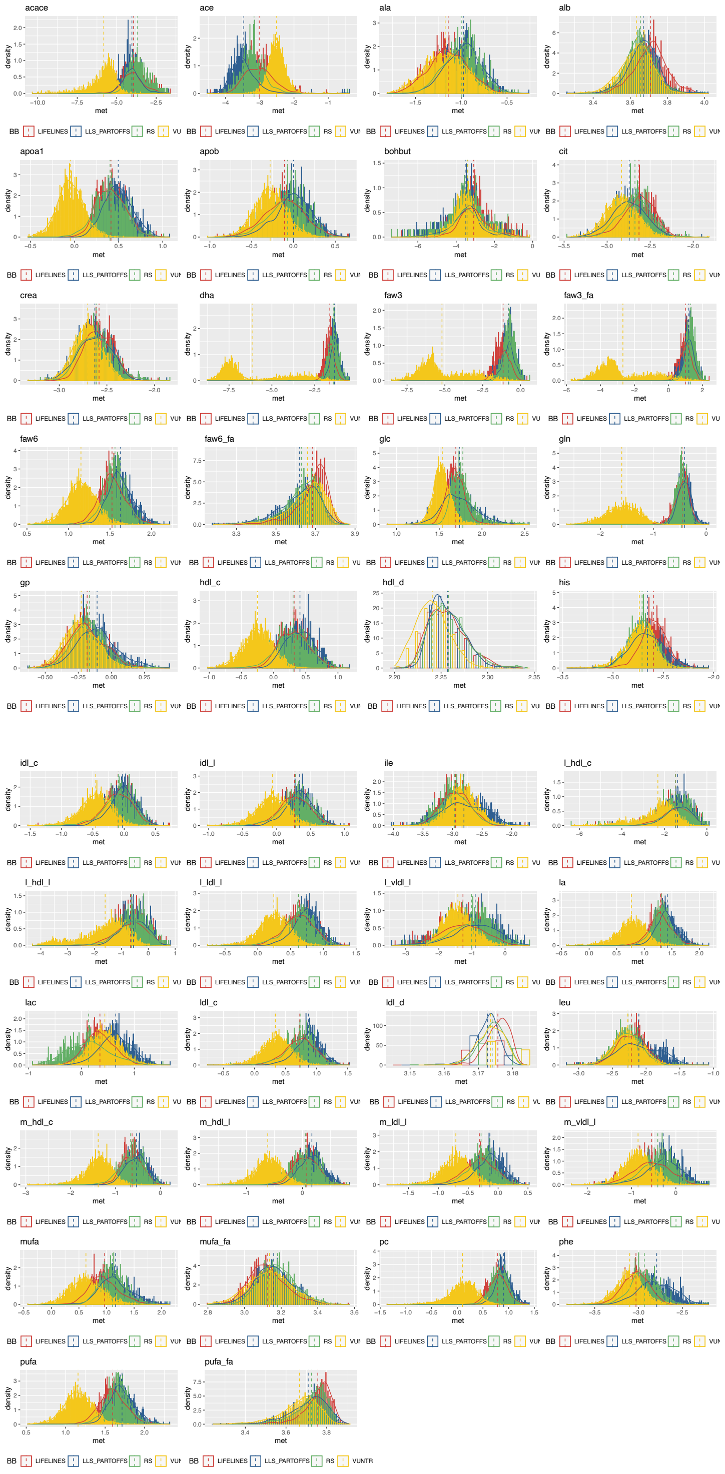

### Distributions after calibrations

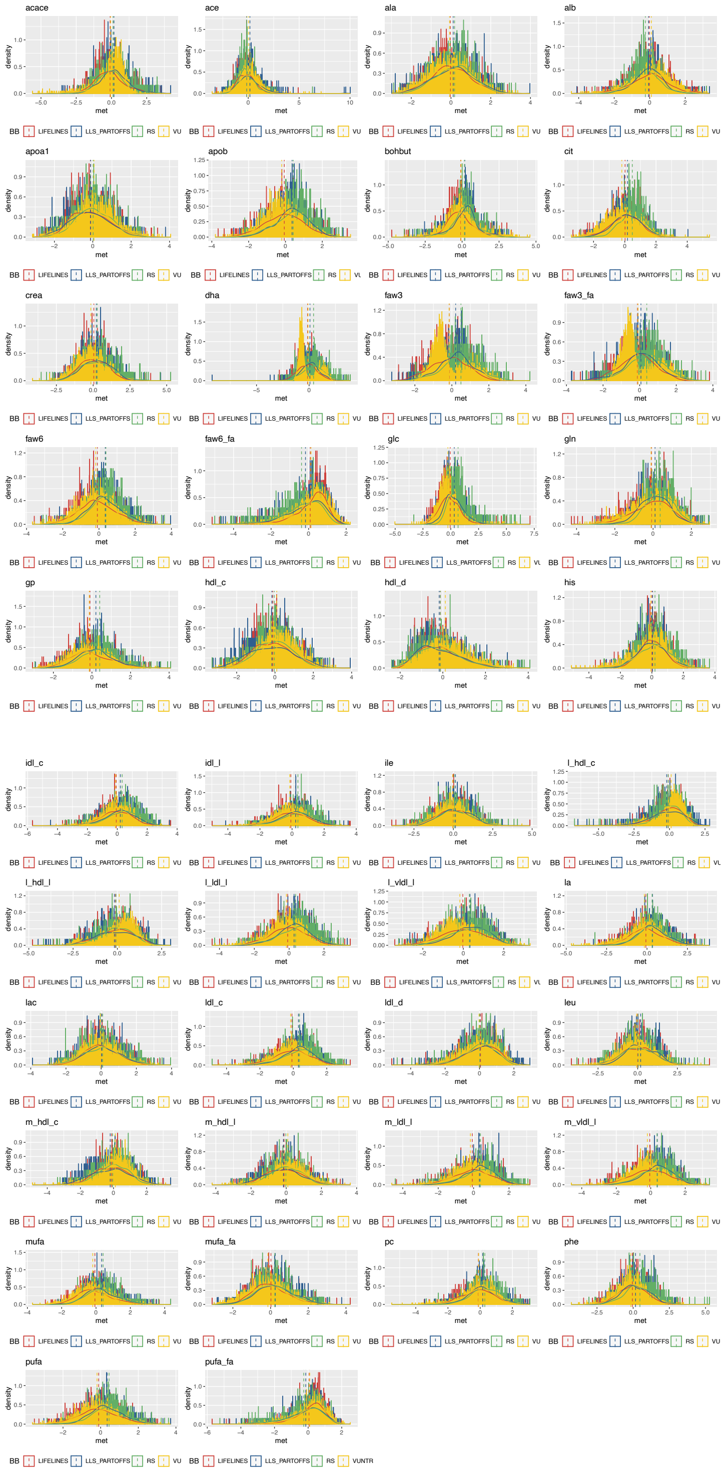

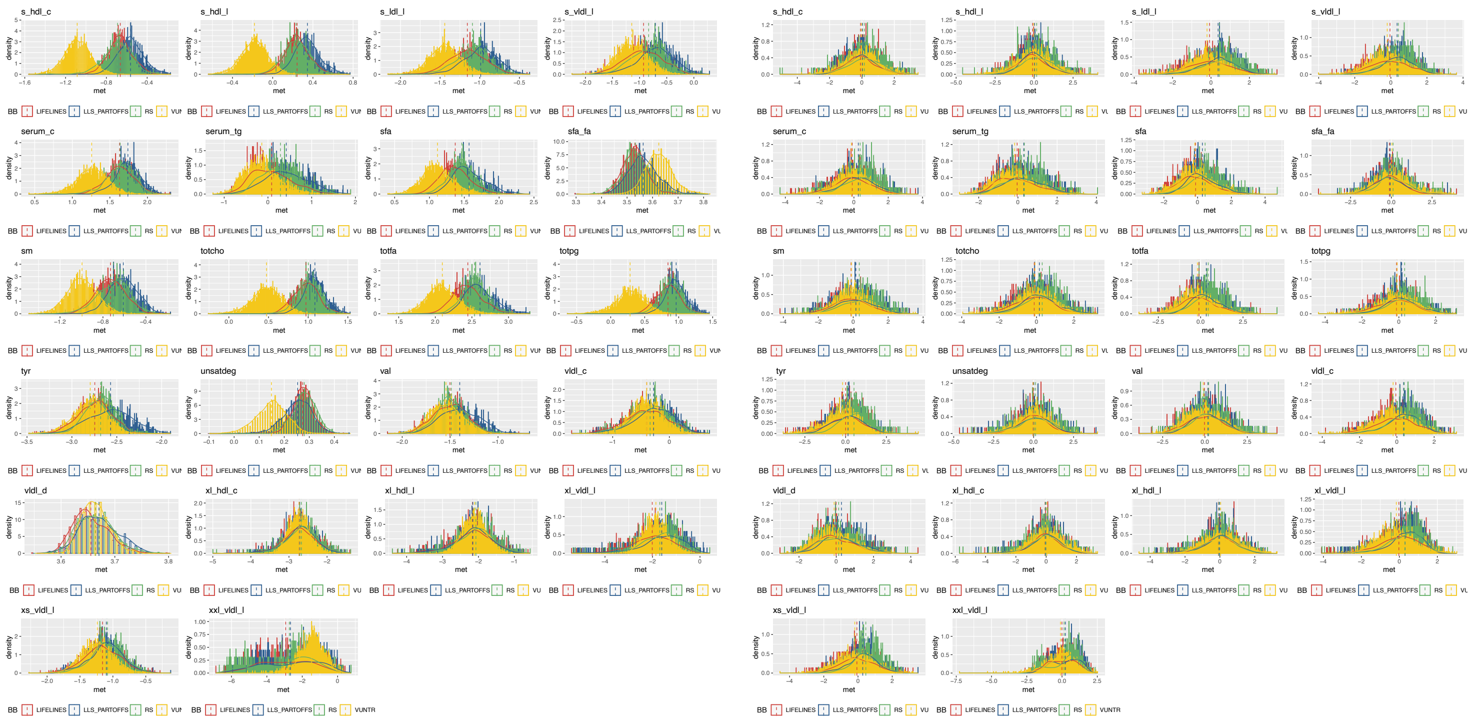

*Figure SM2: Comparisons distributions of each metabolomics feature before (left column) and after (right column) the harmonization technique.*

#### ElasticNET regression model

The model considered to train each DNAm metabolomics feature is regression models:

$$\widehat{m}_i \sim \beta_0 + \sum_{j=1}^{N} \beta_j c_j + \beta_1 + \text{sentrix position} + \varepsilon_i$$

in which $m_i$ represent one of metabolomics feature of interest, $c_j$ one of the CpG sites selected with the pre-selection, $\beta_j$ the regression coefficient, and $\varepsilon_i$ the normal distributed reconstruction error. The regression coefficients are found by minimizing the ordinary least squares error, with the addition of an elastic net regularization term for the coefficients:

$$\boldsymbol{\beta}(\alpha, \lambda) = argmin_{\beta}(\,(c_i - \hat{c}_i)^2 + \lambda[\alpha\|\boldsymbol{\beta}\|_1 + (1-\alpha)\|\boldsymbol{\beta}\|_2])$$

in which $\lambda$ ($\in (0, \infty)$) represents the "shrinkage parameter" and $\alpha$ ($\in (0,1)$) is the mixing parameter balancing the L1 and L2 norm regularizations. We fixed the mixing parameter $\alpha$ at 0.5 for the predictive models, like previously done by other authors and optimized the shrinkage parameter using an inner 5-Fold Cross-Validation scheme [2,3].

#### Mortality associations

We used several Cox regression models in the Rotterdam Study (N = 1542 with 285 reported deaths) to evaluate the mortality associations of the DNA methylation metabolomics features. First build univariate Cox regression models, corrected by sex and age, to evaluate the univariate associations of each of the 64 DNAm metabolomics feature (DNAm metab$_i$) and the DNAm metaboHealth:

$$\mathrm{H(t)} \sim H_0(t)e^{b_1\, DNAm\ metab_i + b_2\, sex + b_3\, age}$$
$$\mathrm{H(t)} \sim H_0(t)e^{b_1\, \mathrm{DNAm\ MetaboHealth} + b_2\, sex + b_3\, age}$$

where $H_0(t)$ is the baseline hazard at time t, and $\mathrm{H(t)}$ is the predicted Hazard.
We used Benjamini-Hochberg to correct for multiple testing.

As a second step we wanted to evaluate if the DNAm metabolomics features could exert also independent mortality signal. Therefore, we trained a stepwise (backward/farward) multivariate Cox regression models using contemporarily all our DNA methylation metabolomics models.

$$\mathrm{step(H(t)} \sim H_0(t)e^{b_1\, sex + b_2\, age + b_3 DNAm\ metab_3 + \cdots + b_{64} DNAm\ metab_{64}})$$

The model started with all the 64 DNAm metabolomics and then optimized the feature selection running for 1000 steps in which the algorithm could include or exclude the features at each iteration based on the Akaike Information Criterium (AIC).
We then evaluated if our DNAm metabolomics features could also add independent signal also in respect to GrimAge, or to its DNAm components (GrimAge surro).

$$\mathrm{step(H(t)} \sim H_0(t)e^{b_1\, sex + b_2\, age + b_3 DNAm\ metab_3 + \cdots + b_{64} DNAm\ metab_{64} + GrimAge})$$

$$\mathrm{step(H(t)} \sim H_0(t)e^{b_1\, sex + b_2\, age + b_3 DNAm\ metab_3 + \cdots + b_{64} DNAm\ metab_{64} + b_{65} GrimAge\ surro_1 + \cdots + b_{73} GrimAge\ surro_8})$$

Following the same principle, as a final evaluation, we also added the 109-plasma protein EpiScores.

### Supplementary tables

Table S1: Phenotypic characteristics BIOS Consortium. *Table with the phenotypic characteristics of the samples in the BIOS Consortium used to train and evaluate the DNAm models.*

| **Characteristic** | **LIFELINES**, N = 729[1] | **LLS_PARTOFFS**, N = 671[1] | **RS**, N = 640[1] | **VUNTR**, N = 2,294[1] |
|---|---|---|---|---|
| age | 45.58 (35.00, 55.00) | 58.31 (54.00, 63.00) | 67.15 (64.00, 71.00) | 37.58 (28.00, 46.00) |
| sex | | | | |
| female | 423 (58%) | 364 (54%) | 369 (58%) | 1,567 (68%) |
| male | 306 (42%) | 307 (46%) | 271 (42%) | 727 (32%) |
| BMI | 25.36 (22.47, 27.44) | 25.42 (23.04, 27.31) | 27.70 (24.80, 29.96) | 24.33 (21.55, 26.29) |
| (Missing) | 0 | 68 | 0 | 8 |

[1] Mean (IQR); n (%)

Table S2: Phenotypic characteristics RS. *Table with the phenotypic characteristics of the samples in the Rotterdam Study used for the mortality analyses.*

| **Characteristic** | **Overall**, N = 1,544[1] | **with metabolomics**, N = 666[1] | **without metabolomics**, N = 878[1] |
|---|---|---|---|
| Age | 64.26 (57.61, 70.60) | 68.01 (65.53, 71.76) | 61.41 (54.69, 65.88) |
| Sex | | | |
| female | 687 (44%) | 285 (43%) | 402 (46%) |
| male | 857 (56%) | 381 (57%) | 476 (54%) |
| Death | | | |
| 0 | 1,259 (82%) | 562 (84%) | 697 (79%) |
| 1 | 285 (18%) | 104 (16%) | 181 (21%) |
| FU_age | 74.66 (68.48, 80.47) | 77.44 (73.93, 81.66) | 72.55 (66.31, 76.34) |
| (Missing) | 2 | 1 | 1 |

[1] Mean (IQR); n (%)

Table S3: Description of the metabolomics variables used to build the predictors.

| name | BBMRI_names | description |
|---|---|---|
| Total-C | serum_c | Total cholesterol |
| VLDL-C | vldl_c | VLDL cholesterol |
| LDL-C | ldl_c | LDL cholesterol |
| HDL-C | hdl_c | HDL cholesterol |
| Total triglycerides | serum_tg | Total triglycerides |
| VLDL particle size | vldl_d | Average diameter for VLDL particles |
| LDL particle size | ldl_d | Average diameter for LDL particles |
| HDL particle size | hdl_d | Average diameter for HDL particles |
| Phosphoglycerides | totpg | Phosphoglycerides |
| Total cholines | totcho | Total cholines |
| Phosphatidylcholines | pc | Phosphatidylcholines |
| Sphingomyelins | sm | Sphingomyelins |
| ApoB | apob | Apolipoprotein B |
| ApoA1 | apoa1 | Apolipoprotein A1 |
| Total fatty acids | totfa | Total fatty acids |
| Unsaturation | unsatdeg | Degree of unsaturation |
| Omega-3 | faw3 | Omega-3 fatty acids |
| Omega-6 | faw6 | Omega-6 fatty acids |
| PUFA | pufa | Polyunsaturated fatty acids |
| MUFA | mufa | Monounsaturated fatty acids |
| SFA | sfa | Saturated fatty acids |
| LA | la | Linoleic acid |
| DHA | dha | Docosahexaenoic acid |
| Omega-3 % | faw3_fa | Ratio of omega-3 fatty acids to total fatty acids |
| Omega-6 % | faw6_fa | Ratio of omega-6 fatty acids to total fatty acids |
| PUFA % | pufa_fa | Ratio of polyunsaturated fatty acids to total fatty acids |
| MUFA % | mufa_fa | Ratio of monounsaturated fatty acids to total fatty acids |
| SFA % | sfa_fa | Ratio of saturated fatty acids to total fatty acids |
| Alanine | ala | Alanine |
| Glutamine | gln | Glutamine |
| Histidine | his | Histidine |
| Isoleucine | ile | Isoleucine |
| Leucine | leu | Leucine |
| Valine | val | Valine |
| Phenylalanine | phe | Phenylalanine |
| Tyrosine | tyr | Tyrosine |
| Glucose | glc | Glucose |
| Lactate | lac | Lactate |
| Pyruvate | pyr | Pyruvate |
| Citrate | cit | Citrate |
| 3-Hydroxybutyrate | bohbut | 3-Hydroxybutyrate |
| Acetate | ace | Acetate |
| Acetoacetate | acace | Acetoacetate |

| | | |
|---|---|---|
| Creatinine | crea | Creatinine |
| Albumin | alb | Albumin |
| Glycoprotein acetyls | gp | Glycoprotein acetyls |
| XXL-VLDL-L | xxl_vldl_l | Total lipids in chylomicrons and extremely large VLDL |
| XL-VLDL-L | xl_vldl_l | Total lipids in very large VLDL |
| L-VLDL-L | l_vldl_l | Total lipids in large VLDL |
| M-VLDL-L | m_vldl_l | Total lipids in medium VLDL |
| S-VLDL-L | s_vldl_l | Total lipids in small VLDL |
| XS-VLDL-L | xs_vldl_l | Total lipids in very small VLDL |
| IDL-L | idl_l | Total lipids in IDL |
| IDL-C | idl_c | Cholesterol in IDL |
| L-LDL-L | l_ldl_l | Total lipids in large LDL |
| M-LDL-L | m_ldl_l | Total lipids in medium LDL |
| S-LDL-L | s_ldl_l | Total lipids in small LDL |
| XL-HDL-L | xl_hdl_l | Total lipids in very large HDL |
| XL-HDL-C | xl_hdl_c | Cholesterol in very large HDL |
| L-HDL-L | l_hdl_l | Total lipids in large HDL |
| L-HDL-C | l_hdl_c | Cholesterol in large HDL |
| M-HDL-L | m_hdl_l | Total lipids in medium HDL |
| M-HDL-C | m_hdl_c | Cholesterol in medium HDL |
| S-HDL-L | s_hdl_l | Total lipids in small HDL |
| S-HDL-C | s_hdl_c | Cholesterol in small HDL |

### BIOS Consortium

#### Consortium banner

#### Cohort Description

##### Leiden Longevity Study (LLS)

The Leiden Longevity Study (LLS) consists of 421 long-lived families of European descent. Families were included if at least two long-lived siblings were alive and fulfilled the age criterion of 89 years or older for males and 91 years or older for females, representing <0·5% of the Dutch population in 2001 [1]. In total, 944 long-lived proband siblings (mean age = 94 years, range = 89-104), 1671 offspring (mean age = 61 years, range = 39-81) and 744 spouses thereof (mean age = 60 years, range = 36-79) were included. Registry-based follow-up until the 27th of October 2016 was available for all participants. Metabolites were successfully quantified in 843 nonagenarians [LLS_SIBS], 1157 of their offspring and 684 controls (LLS_PAROFFS) using non-fasted EDTA plasma samples.

1. M. Schoenmaker et al. Evidence of genetic enrichment for exceptional survival using a family approach: the Leiden Longevity Study. Eur J Hum Genet. 2006 Jan;14(1):79-84.

##### LIFELINES-DEEP

The LifeLines-DEEP cohort is a subset of the Dutch general population cohort LifeLines. Both LifeLines and LifeLines DEEP have been previously described [1-3]. In summary, LifeLines is a three-generation observational follow-up study, which was set up to investigate universal risk factors and their modifiers for multifactorial diseases. Since 2006, approximately 167,000 individuals from the general population residing in the three northern provinces of the Netherlands participate in the study. All participants will be followed-up prospectively for at least 30 years. Participants regularly undergo physical examinations and fill in extensive questionnaires. In addition, blood and urine samples are collected. Each participant is asked to fill in health, lifestyle, and quality-of-life questionnaires every 1.5 years, whereas each participant is invited for a follow-up visit to a Lifelines clinic every 5 years [1,2]. LifeLines-DEEP comprises 1,539 participants (636 males and 903 females, age range 18–84 years). This study was set up for the more detailed phenotyping and omics profiling. For analysis of the genome, epigenome, transcriptome, microbiome, metabolome and other biological levels, additional biomaterials were collected, including additional blood, exhaled air and fecal samples, as well as responses to gastrointestinal health [3].

The current metabolomics study included the baseline information and plasma samples of LifeLines-DEEP participants. EDTA plasma samples were collected after overnight fasting. Peripheral blood samples were drawn by venipuncture from the median cubital vein and subsequently placed at 4°C. Transport of the samples from the research site to the LifeLines laboratory in Groningen was under tightly controlled and continuously monitored conditions. At the LifeLines site, plasma was prepared and aliquoted and stored at -80°C. The samples underwent two freeze-thaw cycles prior to shipment to Brainshake for metabolome analysis [1]. With some sample drop-off, this study eventually included 1,440 LifeLines-DEEP participants. The LifeLines DEEP study was approved by the institutional ethics review board of University Medical Center Groningen (ref. M12.113965).

**Website:** https://www.lifelines.nl/

1. Scholtens, S. et al. Cohort Profile: LifeLines, a three-generation cohort study and biobank. Int. J. Epidemiol. 44, 1172–1180 (2015).
2. Stolk, R. P. et al. Universal risk factors for multifactorial diseases: LifeLines: A three-generation population-based study. Eur. J. Epidemiol. 23, 67–74 (2008).
3. Tigchelaar E. F. et al., Cohort profile: LifeLines DEEP, a prospective, general population cohort study in the northern Netherlands: study design and baseline characteristics. BMJ Open 5, e006772 (2015).

##### Rotterdam Study (RS)

The Rotterdam Study is a prospective, population-based cohort study among individuals living in the well-defined Ommoord district in the city of Rotterdam in The Netherlands [1]. The aim of the study is to determine the occurrence of cardiovascular, neurological, ophthalmic, endocrine, hepatic, respiratory, locomotor, dermatological, otolaryngological, and psychiatric diseases in elderly people. The cohort was initially defined in 1990 among approximately 7,983 persons, aged 55 years and older, who underwent a home interview and extensive physical examination at the baseline and during follow-up visits every 3-4 years (RS-I)[1]. Cohort was extended in 2000/2001 (RS-II, 3,011 individuals aged 55 years and older) and 2006/2008 (RS-III, 3,932 subjects, aged 45 and older). As of 2008, Rotterdam Study comprised 14,926 subjects. Written informed consent was obtained from all participants and the Medical Ethics Committee of the Erasmus Medical Center, Rotterdam, approved the study. Metabolomics measurements were quantified in fasted EDTA plasma samples using Nightingale Health platform. Metabolomics measurements were available for 2,986 participants from RS-I, 591 participants from RS-II (n=591) and 1,787 participants from RS-III.

1. Ikram, M. Arfan, et al. "The Rotterdam Study: 2018 update on objectives, design and main results." *European Journal of Epidemiology* 32.9 (2017): 807-850.

#### NTR

Since 1987, the Netherlands Twin Register is collecting (longitudinal) data in young and adult twins and their families [1,2]. The rich phenotypic longitudinal information that has been collected extends from lifestyle, exposures, personality and demographics information to mental and somatic health. In subgroups information on autonomic and central nervous system function, biomarkers and gene expression, epigenetics and genotyping is available. A 2015 estimate is that, since initiating the NTR, ~25% of all twins and multiples in the Netherlands participated in NTR research projects. Longitudinal information for over 200,000 participants (twins, multiples and family members) was collected over multiple NTR research projects. Data collection is ongoing. A pdf of nearly all published papers may be found at the NTR website.

As part of a Netherlands Twin Register (NTR) biobank project (BB1), 9,530 participants from 3,477 families were visited at home between January 2004 and July 2008 for collection of blood samples [3]. A second project (BB2) collected blood samples in 517 subjects from January 2011 to December 2011, including 210 MZ twin pairs and 64 twin-spouse pairs [4]. Visits were scheduled between 7:00 and 10:00 am and fertile women were bled on day 2-4 of the menstrual cycle, or in their pill-free week. Body composition was measured and information about physical health and lifestyle (e.g. smoking and drinking behavior, exercise, medication use) was obtained. For more detailed information about the methodology of the NTR Biobank study, see [3]. The NTR studies were approved by the Central Ethics Committee on Research involving human subjects of the VUMC, Amsterdam, an Institutional Review Board certified by the US Office of Human Research Protections (IRB number IRB-2991 under Federal wide Assurance-3703; IRB/institute codes, NTR 03-180). All subjects provided written informed consent. Subject were selected who were part of NTR biobank and who in general had rich phenotyping data available.

**website:** www.tweelingenregister.org/

1. Van Beijsterveldt CE, Groen-Blokhuis M, Hottenga JJ, Franić S, Hudziak JJ, Lamb D, Huppertz C, de Zeeuw E, Nivard M, Schutte N, Swagerman S, Glasner T, van Fulpen M, Brouwer C, Stroet T, Nowotny D, Ehli EA, Davies GE, Scheet P, Orlebeke JF, Kan KJ, Smit D, Dolan CV, Middeldorp CM, de Geus EJ, Bartels M, Boomsma DI. The Young Netherlands Twin Register (YNTR): longitudinal twin and family studies in over 70,000 children. *Twin Res Hum Genet.* **2013** Feb; 16(1): 252-267. doi: 10.1017/thg.2012.118. PMID: 23186620
2. Willemsen G, Vink JM, Abdellaoui A, den Braber A, van Beek JH, Draisma HH, van Dongen J, van 't Ent D, Geels LM, van Lien R, Ligthart L, Kattenberg M, Mbarek H, de Moor MH, Neijts M, Pool R, Stroo N, Kluft C, Suchiman HE, Slagboom PE, de Geus EJ, Boomsma DI.The Adult Netherlands Twin Register: twenty-five years of survey and biological data collection.*Twin Res Hum Genet.* **2013** Feb; 16(1): 271-281. doi: 10.1017/thg.2012.140. PMID: 23298648. PMCID: PMC3739974.
3. Willemsen G, de Geus EJ, Bartels M, van Beijsterveldt CE, Brooks AI, Estourgie-van Burk GF, Fugman DA, Hoekstra C, Hottenga JJ, Kluft K, Meijer P, Montgomery GW, Rizzu P, Sondervan D, Smit AB, Spijker S, Suchiman HE, Tischfield JA, Lehner T, Slagboom PE, Boomsma DI. The Netherlands Twin Register biobank: a resource for genetic epidemiological studies. *Twin Res Hum Genet.* **2010** Jun; 13(3): 231-245. doi: 10.1375/twin.13.3.231. PMID: 20477721.
4. Sirota M, Willemsen G, Sundar P, Pitts SJ, Potluri S, Prifti E, Kennedy S, Ehrlich SD, Neuteboom J, Kluft C, Malone KE, Cox DR, de Geus EJ, Boomsma DI. Effect of genome and environment on metabolic and inflammatory profiles. *PLoS One.* **2015** Apr 8; 10(4): e012089. doi: 10.1371/journal.pone.0120898. PMID: 25853885. PMCID: PMC4390246.
5 van Dongen J, Nivard MG, Willemsen G, Hottenga JJ, Helmer Q, Dolan CV, Ehli EA, Davies GE, van Iterson M, Breeze CE, Beck S, Bios Consortium, Suchiman HE, Jansen R, van Meurs JB, Heijmans BT, Slagboom PE, Boomsma DI. Genetic and environmental influences interact with age and sex in shaping the human methylome. Nature Communication. 2016 Apr 7; 7(1): 11115. doi: 10.1038/ncomms11115 (2016). PMID: 27051996. PMCID: PMC4820861.

#### Acknowledgements

##### Leiden Longevity Study (LLS)

The LLS has received funding from the European Union's Seventh Framework Programme (FP7/2007-2011) under grant agreement n° 259679. This study was supported by a grant from the Innovation-Oriented Research Program on Genomics (SenterNovem IGE05007), the Centre for Medical Systems Biology, and the Netherlands Consortium for Healthy Ageing (grants 05040202 and 050-060-810), all in the framework of the Netherlands Genomics Initiative, Netherlands Organization for Scientific Research (NWO), Unilever Colworth, and by BBMRI-NL, a Research Infrastructure financed by the Dutch government (NWO 184.021.007 and 184.033.111).

##### LIFELINES-DEEP

We thank the participants and staff of the Lifelines cohort for their collaboration, particularly B. Bolmer and S. Gerritsma for coordinating the Lifelines data. We thank J. Dekens and J. Arends for management and technical support and the Genomics Coordination Center for providing data infrastructure and access to high performance computing clusters. This project was funded by the Netherlands Heart Foundation (IN-CONTROL CVON grant 2012-03 and 2018-27), the grant from the Top Institute Food and Nitrition (TiFN GH001), the Netherlands Organization for Scientific Research (NWO) 024.004.017, 864.13.013, 016.178.056,

917.14.374, SPI 92-266; the European Research Council Consolidate grant 101001678; and the RuG Investment Agenda Grant Personalized Health grant.

#### Rotterdam Study (RS)

The Rotterdam Study is supported by the Erasmus MC University Medical Center and Erasmus University Rotterdam; The Netherlands Organisation for Scientific Research (NWO); The Netherlands Organisation for Health Research and Development (ZonMw); the Research Institute for Diseases in the Elderly (RIDE); The Netherlands Genomics Initiative (NGI); the Ministry of Education, Culture and Science; the Ministry of Health, Welfare and Sports; the European Commission (DG XII); and the Municipality of Rotterdam. The authors are grateful to the study participants, the staff from the Rotterdam Study and the participating general practitioners and pharmacists. Metabolomics measurements were funded by Biobanking and Biomolecular Resources Research Infrastructure (BBMRI)–NL (184.021.007) and the JNPD under the project PERADES (grant number 733051021, Defining Genetic, Polygenic and Environmental Risk for Alzheimer's Disease using multiple powerful cohorts, focused Epigenetics and Stem cell metabolomics).

#### VUNTR

Funding was obtained from the Netherlands Organization for Scientific Research (NWO) and MagW/ZonMW grants 904-61-090, 985-10-002, 904-61-193,480-04-004, 400-05-717, Addiction-31160008, Middelgroot-911-09-032, Spinozapremie 56-464-14192, Biobanking and Biomolecular Resources Research Infrastructure (BBMRI –NL, 184.021.007).; the European Community's Seventh Framework Program (FP7/2007-2013), ENGAGE (HEALTH-F4-2007-201413); the European Science Council (ERC Advanced, 230374), Rutgers University Cell and DNA Repository (NIMH U24 MH068457-06), the Avera Institute, Sioux Falls, South Dakota (USA) and the National Institutes of Health (NIH, R01D0042157-01A, MH081802, Grand Opportunity grants 1RC2 MH089951). We gratefully acknowledge grant NWO 480-15-001/674: Netherlands Twin Registry Repository: researching the interplay between genome and environment.

### Ethics statements

#### Leiden Longevity Study (LLS)

The Leiden Longevity Study protocol was approved by the Medical Ethical Committee of the Leiden University Medical Center before the start of the study (P01.113). In accordance with the Declaration of Helsinki, the Leiden Longevity Study obtained informed consent from all participants prior to their entering the study.

#### LIFELINES-DEEP

The LifeLines DEEP study was approved by the ethics committee of the University Medical Center Groningen, document number METC UMCG LLDEEP: M12.113965. All participants signed an informed consent form before study enrollment. All procedures performed in studies involving human participants were in accordance with the ethical standards of the institutional and/or national research committee and with the 1964 Helsinki declaration and its later amendments or comparable ethical standards.

#### Rotterdam Study

*The Rotterdam S*tudy protocol was approved by the Medical Ethics Committee of the Erasmus MC Rotterdam, the Nethrlands. (*MEC 02.1015*) and by the Dutch Ministry of Health, Welfare, and Sport (Population Screening Act WBO, license number 1071272-159521-PG). In accordance with the Declaration of Helsinki, the *Rotterdam Study* obtained written informed consent from all participants prior to their entering the study.

#### VUNTR

The Netherlands twin Register study protocol was approved by the Central Ethics Committee on Research Involving Human Subjects of the VU University Medical Centre, Amsterdam, an Institutional Review Board certified by the U.S. Office of Human Research Protections (IRB number IRB00002991 under Federal-wide Assurance- FWA00017598. In accordance with the Declaration of Helsinki, the Netherlands Twin Register obtained informed consent from all participants prior to their entering the study.
